# GLP-1 Receptor Agonists (GLP-1RAs) for treatment of tobacco and alcohol dependence: a systematic review and meta-analysis of randomised controlled trials

**DOI:** 10.64898/2026.10.21.26366084

**Authors:** Gemma Taylor, Chloe Burke, Daniel Titheradge, Shiying Jia, Beyza Cihan, Isabel Faulkner, Gareth Williams, Kushala Abeysekera, Matthew Hickman, Kyla Thomas

## Abstract

**Background:** Glucagon-like peptide-1 receptor agonists (GLP-1RAs) might reduce tobacco and alcohol use. We synthesised evidence from randomised controlled trials (RCTs) on smoking and alcohol outcomes, including secondary outcomes reported in trials conducted for other clinical purposes.

**Methods:** We searched MEDLINE, Embase, PsycINFO, Web of Science, CENTRAL, PubMed, and registries for RCTs comparing GLP-1RAs with placebo from inception to August 18, 2026. Smoking and alcohol outcomes were synthesised using random-effects meta-analyses. We assessed risk of bias (RoB) with RoB-2, and certainty of evidence with the Grading of Recommendations Assessment, Development and Evaluation (GRADE) approach.

**Findings:** Fourteen trials involving 1149 participants were included; 13 contributed to meta-analyses. Across the 14 trials, 8 recruited participants not seeking either tobacco or alcohol treatment. The other 6 recruited treatment-seeking participants: 3 for tobacco and 3 for alcohol. Eight trials reported smoking-related outcomes, 11 reported alcohol outcomes, and 5 reported both. Follow-up ranged from 6 to 60 weeks.

All 4 pooled smoking-related estimates favoured GLP-1RAs, although all 95% CIs included no difference and were compatible with increases and decreases in outcomes; e.g., smoking cessation, OR 1.39 (95% CI, 0.77 to 2.53, k=3). Restricting analyses to semaglutide trials indicated a reduction in cigarettes per day (CPD) (standardised mean difference [SMD]=-0.10, 95%CI:-0.16 to -0.04; k=3).

Across agents, pooled estimates favoured GLP-1RAs for days without alcohol (SMD=0.35, 95%CI: 0.12 to 0.59, k=3). Pooled estimates of semaglutide-only trials also favoured treatment for alcohol consumption (grams/day; SMD=-0.56, 95%CI:-0.82 to -0.30; k=2), craving (SMD=-0.60, 95%CI:-0.91 to -0.29; k=2), drinks per drinking day (SMD=-0.59, 95%CI:-0.86 to -0.32; k=3), drinks per calendar day (SMD=-0.48, 95%CI:-0.87 to -0.09; k=2) and heavy drinking days (SMD=-0.45, 95%CI:-0.85 to -0.06; k=3), and alcohol-free days (SMD=0.41, 95%CI:0.09 to 0.73; k=2).

Two of 41 assessed results were at low RoB, 31 raised some concerns and 8 were at high risk; certainty was moderate for 1 outcome, low for 4 and very low for 8.

**Interpretation:** GLP-1RAs may reduce alcohol consumption and CPD, with the clearest evidence for semaglutide. These findings should be contextualised by most evidence being low- to very-low-certainty. Larger trials are needed to establish clinically meaningful benefit and sustained abstinence.

**KEY POINTS:** *Question:* Do glucagon-like peptide-1 receptor agonists improve smoking and alcohol outcomes compared with placebo?

*Findings:* This systematic review included 14 randomised trials involving 1149 participants, with 13 trials contributing to meta-analyses. Alcohol-free days increased significantly across agents, and semaglutide analyses indicated significant reductions in alcohol consumption; and cigarettes per day. There was insufficient evidence on smoking-cessation result.

*Meaning:* GLP-1 receptor agonists show potential for reducing alcohol consumption, and cigarettes per day, but predominantly low or very low certainty evidence warrants larger trials before their role in addiction treatment can be established.

## INTRODUCTION

Smoking and alcohol use account for more than 7.7^1^ and 2.6 million^2^ deaths annually, with estimated global costs of USD$1.4 and $2.6 trillion, respectively^3,4^. These behaviours frequently co-occur, disproportionately affecting people with mental health conditions and socioeconomic disadvantage^5,6^, and contribute to preventable multimorbidity and premature mortality^7-11^. Yet both remain under-addressed in routine healthcare, particularly among underserved groups^12,13^.

Although smoking cessation substantially improves physical and mental health^14,15^, concerns about post-cessation weight gain can deter quit attempts^16^, and no available treatment reliably prevents this weight gain^16^. Existing cessation medicines have limitations in effectiveness and are cautioned among some groups, including people with type 2 diabetes (T2D) and mental health conditions^12,13,17,18^. Pharmacotherapies for alcohol use disorder (AUD), including acamprosate, naltrexone and disulfiram, have modest effects, low uptake and adherence, and relapse is high with slightly stronger evidence for maintaining abstinence in people with severe AUD^19-22^.

Glucagon-like peptide-1 receptor agonists (GLP-1RAs), licensed for T2D and obesity, regulate appetite and influence mesolimbic dopaminergic reward pathways implicated in tobacco and alcohol addiction^23^. In animal models, they reduce nicotine self-administration and withdrawal, alcohol consumption, conditioned place preference and relapse-like behaviour^24,25^. Observational studies have reported greater engagement with tobacco-treatment services and reduced alcohol consumption among people prescribed GLP-1RAs^26^, while observational meta-analyses suggest reductions in alcohol consumption, alcohol-use disorder (AUD) diagnoses and Alcohol Use Disorders Identification Test (AUDIT) scores^27,28^. However, these findings remain vulnerable to confounding and selection bias.

Randomised controlled trial (RCT) evidence is limited and mixed. Dulaglutide added to varenicline did not improve smoking cessation but reduced post-cessation weight gain^29^, whereas exenatide combined with nicotine replacement therapy increased quit rates and reduced weight gain^30^. Trials are small, underpowered and clinically heterogeneous, and meta-analyses have found smaller, uncertain effects on alcohol consumption and craving^27,31-33^. Existing reviews have focused largely on alcohol or observational evidence, with tobacco outcomes synthesised narratively^27,28,31,32,34-36^. Some did not include eligible trials available at the time of searching^27,32,33^, while one appeared to pool baseline rather than outcome data and may have assessed risk of bias (RoB) too favourably^36^.

### Aims

To synthesise GLP-1RA RCTs evaluating smoking- and alcohol-related outcomes.

## METHODS

We conducted a PROSPERO-registered systematic review and meta-analysis (CRD420251155420), following PRISMA guidance. Protocol deviations are reported in the supplement. Manuscript pre-print available via medRxiv (MEDRXIV/2026/366084), and code available via GitHub (insert URL upon publication).

### Eligibility Criteria

We included RCTs comparing GLP-1RAs or dual agonists with any comparator in people who smoked or consumed alcohol, reporting cessation, consumption, craving, dependence or risk outcomes (eTable-1). We selected the longest outcome-specific follow-up of ≥28 days with sufficient data for analysis, which could precede the trial’s final follow-up.

### Search Methods, Data Collection and Analysis

We searched MEDLINE, Embase, PsycINFO, Web of Science, CENTRAL, PubMed, and trial registries from inception to August 18, 2026, and searched relevant reviews’ reference lists. Search strategies are presented in the supplement.

Using Covidence, two reviewers independently screened titles, abstracts and full texts, extracted outcome data and assessed RoB; disagreements were resolved through discussion or third-reviewer adjudication. One reviewer extracted study characteristics using a piloted form, checked by a second reviewer. We extracted estimates and variances or group-level data sufficient to calculate standardised mean differences (SMDs) or odds ratios (ORs). We assessed Risk of Bias (RoB) using the Cochrane RoB-2 tool^37^, and certainty of evidence for primary outcomes using the Grading of Recommendations Assessment, Development and Evaluation (GRADE) approach^38^.

We conducted separate random-effects meta-analyses of clinically comparable smoking and alcohol outcomes using Stata’s *meta* package^39^. Random-effects models used restricted maximum likelihood to estimate between-study variance, with normal-based confidence intervals (CIs) and two-sided z-tests for pooled effects. Continuous treatment effects were expressed as Hedges *g* SMDs with 95%CIs. We prioritised adjusted between-group estimates, followed by differences in change scores and baseline and final values. To ensure comparable standardisation, each treatment contrast was divided by the pooled within-group baseline SD, with the Hedges small-sample correction applied. Where required, change-score variances were calculated using established methods^39,40^ and an assumed within-participant correlation of 0.59^14,40-43^. Dichotomous outcomes were expressed as ORs with 95%CIs, prioritising reported adjusted estimates, convertible effect estimates and then event counts. Heterogeneity was assessed using I^2^, χ^2^ and forest plots. Exploratory sensitivity analyses examined GLP-1RA agents (i.e., restricted to the GLP-1RA most frequently represented among trials contributing to each outcome), AUD diagnosis and alcohol or tobacco treatment-seeking status and excluded high-RoB trials. Funnel plots and Egger’s tests were exploratory because all meta-analyses included <10 trials; Egger’s tests were not conducted for outcomes with only 2 trials^41^.

## RESULTS

### Study Selection and Characteristics

We identified 12583 records, screened 4333 titles and abstracts, and assessed 298 full-text reports (Figure-1). Fourteen RCTs involving 1149 participants met the inclusion criteria^29,30,44-55^. Characteristics and associated reports are presented in Table-1, and eTables-2-4; examples of excluded reports are listed in eTable 5. Thirteen trials, with 1124 participants, contributed to at least one meta-analysis; Fogelman et al was synthesised narratively because sufficient numerical data were unavailable^46^. Trial authors were contacted about missing or unclear data (eTable-6).

Across the 14 trials, 8 recruited participants not seeking either tobacco or alcohol treatment^44,46-48,51-53,55^. The other 6 recruited treatment-seeking participants: 3 for tobacco^29,30,45^ and 3 for alcohol^49,50,54^. No trials required detox from alcohol. Trials were conducted in Denmark (n=5)^49-51,53,55^, the US (n=6)^30,45-48,54^, the UK (n=2)^44,52^, and Switzerland (n=1)^29^. Populations included people with metabolic conditions or elevated metabolic risk, smokers with varying treatment intentions, people with AUD, and those with schizophrenia. Ten trials recruited participants with metabolic conditions or elevated metabolic risk^30,44-46,50-55^. Mean age ranged from 35.0 to 57.2 years, and female representation from 30.5% to 100%. Six trials received industry funding or support^44,45,51-53,55^, and industry-related author conflicts were common^29,44,47-55^. Data were obtained primarily from peer-reviewed publications, with 1 trial available only through a registry^45^ and 1 as a conference report^46^.

### Interventions and Comparators

Five trials evaluated liraglutide^44,45,51,52,55^, six semaglutide^46-48,50,53,54^, two exenatide^30,49^, and one dulaglutide^29^. Treatment lasted 6 to 52 weeks and varied in dose and route; all but 1 intervention was administered subcutaneously^54^. All comparator groups received placebo. Behavioural support varied from no addiction-focused treatment to structured smoking-cessation counselling^29,30,45^, or AUD treatment^49,50^. Where provided, support was equivalent between groups.

### Outcomes

Eight trials reported smoking-related outcomes^29,30,45,47-50,53^, 11 reported alcohol outcomes^29,44,46,47,49-55^, and 5 reported both^29,47,49,50,53^. Follow-up ranged from 6 to 60 weeks. Three trials reported biochemically verified smoking abstinence^29,30,45^ and 1 measured phosphatidylethanol^49^; other outcomes were primarily self-reported.

### Ongoing Trials

Thirty ongoing or completed trials awaiting results are described in e-Table-4.

### Effects of Interventions

Smoking and alcohol meta-analyses are shown in Figures-2 and -3, respectively; estimates by analysis are summarised in Table-2.

#### Smoking Cessation

Three trials evaluated smoking cessation with liraglutide, dulaglutide, or exenatide^29,30,45^, in tobacco-treatment-seeking populations. The pooled estimate was in the direction of benefit with treatment, although the 95%CI included no difference, and lower or higher odds of cessation (OR=1.39; 95%CI:0.77 to 2.53; p=0.28; k=3, n=377). Between-study heterogeneity was low (I^2^=23.5%; τ^2^=0.08).

Excluding 1 high RoB trial^45^, produced an OR=1.46 (95%CI:0.70 to 3.03, k=2, n=337).

#### Cigarettes Smoked per Day (CPD)

Four trials evaluated CPD: 3 with semaglutide and 1 with dulaglutide^29,47,48,50^. One recruited participants seeking tobacco treatment^29^; 3 CPD results were at high RoB^47,48,50^. The pooled estimate favoured GLP-1RAs, but the 95%CI included increases and decreases in CPD (SMD=-0.21; 95%CI:-0.44 to 0.02; p=0.08, k=4, n=400). Heterogeneity was moderate (I^2^=61.0%; τ^2^=0.03).

Restricting the analysis to semaglutide trials favoured treatment (SMD=-0.10; 95%CI:-0.16 to -0.04; P<0.001; I^2^=0%; τ^2^=0, k=3, n=145).

#### Nicotine Dependence

Three trials evaluated nicotine dependence using the Fagerström Test for Nicotine Dependence (FTND): 1 with exenatide and 2 with semaglutide. None recruited participants seeking tobacco treatment; all 3 results were at high RoB^49,50,53^. The pooled estimate favoured GLP-1RAs, but the 95%CI included increases and decreases in dependence (SMD=-0.08; 95%CI:-0.60 to 0.44; p=0.76, k=3, n=208), with substantial heterogeneity (I^2^=62.8%; τ^2^=0.13).

In the semaglutide-restricted analysis, the pooled estimate favoured treatment but the CI included no difference and was compatible with either an increase or a decrease in dependence (SMD=-0.28, 95%CI:-1.17 to 0.60; p=0.53; k=2, n=135), with substantial heterogeneity (I^2^=74.4%; τ^2^=0.31).

#### Post-Cessation Weight Gain

Two trials evaluated post-cessation weight gain: 1 with dulaglutide and 1 with exenatide. Both recruited participants seeking tobacco treatment; neither result was at high RoB^29,30^. The pooled estimate favoured GLP-1RAs, but the 95%CI included increases and decreases in post-cessation weight gain (SMD=-0.27, 95%CI:-0.58 to 0.05; p=0.10; k=2, n=104). Heterogeneity was low (I^2^=7.1%; τ^2^=0.01).

#### Alcohol Grams Daily

Four trials evaluated daily alcohol consumption in grams: 1 with liraglutide, 1 with exenatide and 2 with semaglutide^44,47,49,50^. One result was at high RoB^44^. The pooled estimate favoured GLP-1RAs, but the 95%CI included increases and decreases in consumption (SMD=-0.30, 95%CI:-0.63 to 0.04; p=0.08; k=4, n=328). Heterogeneity was substantial (I^2^=61.9%; τ^2^=0.07).

Restricting the analysis to alcohol-treatment seeking populations^49,50^ the pooled estimate favoured GLP-1RAs, but the 95%CI included no difference and was compatible with either an increase or a decrease in consumption (SMD=-0.33, 95%CI:-0.93 to 0.27, k=2, n=235). Restricting the analysis to AUD populations^47,49,50^, the pooled estimate favoured GLP-1RAs (SMD=-0.38, 95%CI:-0.74 to -0.02, k=3, n=283). Excluding one trial with high RoB^44^, the pooled estimate favoured GLP-1RA (SMD=-0.38, 95%CI:-0.74 to -0.02, k=3, n=283).

After excluding the exenatide^49^ and liraglutide trials^44^, the pooled estimate favoured semaglutide (SMD=-0.56, 95%CI:-0.82 to -0.30; p<0.001; k=2, n=156), with no observed heterogeneity (I^2^=0%; τ^2^=0).

#### Alcohol Use Disorders Identification Test (AUDIT)

Five trials contributed AUDIT data: one of exenatide, 2 semaglutide, and 2 liraglutide; none were high RoB^49-53^. The pooled estimate favoured GLP-1RAs, but the 95%CI included increases and decreases in AUDIT scores (SMD=-0.03, 95%CI:-0.29 to 0.23; p=0.82; k=5, n=471). Heterogeneity was substantial (I^2^=65.1%; τ^2^=0.06).

Restricting the analysis to alcohol-treatment seeking populations^49,50^ the pooled estimate favoured GLP-1RAs, but the 95%CI included no difference and was compatible with either an increase or a decrease in AUDIT scores (SMD=-0.23, 95%CI:-0.87 to 0.42,, k=2, n=235). Restriction to AUD populations excluded the same three trials.

There was no evidence that effects differed by GLP-1RA agent (test for subgroup differences: χ^2^=0.50, df=2; p=0.78). Subgroup estimates were SMD=0.10 for exenatide (95%CI:-0.25 to 0.45; k=1, n=127), SMD=0.09 for liraglutide (95%CI:-0.09 to 0.27; k=2, n=163) and SMD=-0.19 for semaglutide (95%CI:-0.94 to 0.57; k=2, n=181).

#### Alcohol Craving

Three trials evaluated alcohol craving using the Penn Alcohol Craving Scale: 1 with exenatide and 2 with semaglutide. All trials recruited participants with AUD seeking alcohol treatment; no results were at high RoB^49,50,54^. The pooled estimate favoured GLP-1RAs but was imprecise with the 95%CI including no effect and compatible with an increase or decrease in craving (SMD=-0.33, 95%CI:-0.85 to 0.18; p=0.20; k=3, n=285). Heterogeneity was considerable (I^2^=77.9%; τ^2^=0.16).

Across the 2 semaglutide trials^50,54^, the pooled estimate favoured treatment (SMD=-0.60, 95%CI:-0.91 to -0.29; I^2^=0%; k=2, n=158).

#### Drinks per Drinking Day

Three semaglutide trials evaluated drinks per drinking day in participants with AUD, and none were at high RoB^47,50,54^. The pooled estimate favoured semaglutide (SMD=-0.59, 95%CI:-0.86 to -0.32; p<0.001; k=3, n=206), with no observed heterogeneity (I^2^=0%; τ^2^=0).

Restricting the analysis to alcohol-treatment seeking populations^50,54^ favoured treatment (SMD=-0.55, 95%CI:-0.86 to -0.24, k=2, n=158).

#### Drinks per Calendar Day

Two semaglutide trials evaluated drinks per calendar day in participants with AUD^47,54^. One recruited participants seeking alcohol treatment and 1 recruited participants not seeking alcohol treatment; both results raised some concerns about bias. The pooled estimate favoured semaglutide (SMD=-0.48, 95%CI:-0.87 to -0.09; p=0.02; k=2, n=98), with no observed heterogeneity (I^2^=0%; τ^2^=0).

#### Drinking Days

Two semaglutide trials reported drinking days in participants with AUD: 1 recruited participants seeking alcohol treatment and 1 recruited participants not seeking alcohol treatment. Both results raised some concerns about risk of bias^47,54^. The pooled estimate favoured semaglutide but the 95%CI included no difference and was compatible with either an increase or a decrease in drinking days (SMD=-0.06, 95%CI:-0.18 to 0.05; p=0.27; k=2, n=98). No heterogeneity was observed (I^2^=0%; τ^2^=0).

#### Drinks per Week

Three trials evaluated drinks per week: 2 with liraglutide and 1 with dulaglutide^29,51,55^. None specifically recruited AUD populations or participants seeking alcohol treatment. All 3 results raised some concerns about bias. The pooled estimate favoured GLP-1RAs, but the 95%CI included increases and decreases in drinks per week (SMD=-0.12, 95%CI:-0.29 to 0.05; p=0.16; k=3, n=330). Heterogeneity was low (I^2^=9.9%; τ^2^=0.01).

After excluding the dulaglutide trial^29^, the pooled estimate favoured placebo but the 95%CI included no difference and was compatible with either an increase or a decrease in drinks per week (SMD=0.04, 95%CI:-0.26 to 0.34; p=0.81; k=2, n=179), with no observed heterogeneity (I^2^=0%; τ^2^=0).

#### Heavy Drinking Days

Four trials contributed data on heavy drinking days: three of semaglutide and 1 exenatide^47,49,50,54^, all trials were of AUD populations, and no trials were high RoB. The pooled estimate favoured GLP-1RAs, but the 95%CI included increases and decreases in heavy drinking days (SMD=-0.28, 95%CI:-0.71 to 0.15; p=0.21; k=4, n=249). Heterogeneity was considerable (I^2^=80.4%; τ^2^=0.15).

Restricting the analysis to trials of alcohol-treatment seeking populations^49,50,54^, the pooled estimate favoured GLP-1RAs, but the 95%CI included increases and decreases in heavy drinking days (SMD=-0.36, 95%CI:-0.95 to 0.24, k=3, n=201).

After excluding the exenatide trial^49^, the pooled estimate favoured semaglutide (SMD=-0.45, 95%CI:-0.85 to -0.06; p=0.03; k=3, n=206), although heterogeneity remained substantial (I^2^=62.5%; τ^2^=0.08).

#### Days without Alcohol Consumption

Three trials contributed data on days without alcohol consumption: two of semaglutide and 1 exenatide; all trials were of AUD populations, and none had high RoB^47,49,50^. The pooled estimate favoured GLP-1RAs (SMD=0.35, 95%CI:0.12 to 0.59; p=0.003; k=3, n=277), with no observed heterogeneity (I^2^=0%; τ^2^=0).

Restricting the analysis to two trials of alcohol-treatment seeking participants^49,50^ the pooled estimate favoured GLP-1RAs (SMD=0.32, 95%CI:0.07 to 0.58, k=2, n=235). Restricting the analysis to 2 semaglutide trials favoured semaglutide (SMD=0.41, 95%CI:0.09 to 0.73; p=0.01; I^2^=0%, k=2, n=150).

### Narrative Review of Alcohol Outcomes

Fogelman et al reported fewer weekly drinking days with semaglutide, but an effect estimate and 95%CI were unavailable^46^ (P<0.045). Semaglutide was also associated with greater reductions in WHO drinking-risk level in Klausen et al^50^ (between-group difference, -0.52; 95%CI:-0.89 to -0.16; p=0.006; number needed to treat=4.3). In Schacht et al^54^, more participants improved by ≥1 risk category with semaglutide (81.0% vs 53.8%; p=0.045), although the finding was attenuated when participants who withdrew were assumed to have made no improvement.

### Risk of Bias

Of 41 assessed results from 13 trials, 2 (4.9%) were at low RoB, 31 (75.6%) raised some concerns, and 8 (19.5%) were at high risk. Most concerns arose from substantial or differential missing outcome data likely related to participants’ alcohol or tobacco use (eFigures-1-3).

### Certainty of Evidence (GRADE)

Certainty was low or very low for most outcomes and moderate for post-cessation weight gain. Downgrading reflected imprecision, within-study bias, particularly missing outcome data, and unexplained heterogeneity (eTables-7-8).

### Small-Study Effects

Funnel plots contained only 2 to 5 trials per outcome and were interpreted descriptively (eFigures-4-5). The FTND test yielded Egger’s test=-16.72, p=0.04, but with only 3 trials this cannot establish publication bias; non-significant tests for other outcomes cannot exclude it (eTable-9).

## DISCUSSION

Across 14 RCTs, pooled estimates generally favoured GLP-1RAs, with clearer evidence for alcohol-consumption than for smoking outcomes. All 4 pooled smoking pooled estimates favoured GLP-1RAs, but their 95% CIs included no difference and remained compatible with a range of effects. The semaglutide-restricted estimate indicated a reduction in cigarettes per day, although 1 trial contributed almost all the evidence. Across agents, the pooled estimates favoured GLP-1RAs showing an increase in alcohol-free days. Restricting analysis to semaglutide-only trials showed reductions in alcohol consumption, alcohol craving, drinks per drinking day, drinks per calendar day and heavy drinking days, and an increase in alcohol-free days.

### Interpretation, Strengths, and Limitations

Consumption may decrease without abstinence, particularly when abstinence is not the treatment goal. Only 3 of 8 smoking trials recruited participants seeking tobacco treatment, and 3 of 11 alcohol trials recruited participants seeking alcohol treatment. Outcome selection may reflect trial purpose. AUDIT may respond less to short-term changes because some items concern earlier events.

Reduced consumption is consistent with modulation of nicotine and alcohol reward, supported by animal studies reporting reduced self-administration^24,25,56^; however, human trials cannot establish this mechanism. Stronger findings for semaglutide may reflect differences in populations, outcomes and trial design; however, superiority cannot be established without direct comparisons. Preliminary animal and observational human evidence also suggest that the dual GIP/GLP-1 receptor agonist tirzepatide may reduce alcohol consumption, warranting trials in people with alcohol use disorder^57,58^.

Sensitivity analyses in treatment-seeking and AUD populations were exploratory and often imprecise; few trials and overlapping populations limited assessment of effect modification. Excluding the high-RoB cessation trial changed the estimate little. RoB restrictions left insufficient trials to assess robustness for CPD and nicotine dependence.

By including secondary tobacco and alcohol-use outcomes, registry results and investigator-provided data, we broadened the evidence base and reduced reliance on published primary-outcome reports, although reporting bias remains possible.

Few trials contributed to each outcome, limiting assessment of small-study effects and subgroup differences and precluding meta-regression. Only 2 of 41 results were at low RoB, and missing outcome data were the principal concern. Trials were conducted in high-income countries, alcohol outcomes were largely self-reported, and populations, treatment duration, support, outcome definitions and follow-up varied. However, adjunct interventions were balanced between groups, providing clear estimates for active versus placebo GLP-1RAs. Industry funding or conflicts were common and may be associated with favourable findings^59^. Imprecision principally limited certainty; and most analyses did not meet the optimal information size.

Clinical importance depends on whether reduced consumption lowers harm and meets patients’ treatment goals. Standardised effects cannot be read directly as drinks or cigarettes avoided; reporting absolute changes would improve interpretation across consumption levels and AUD severity.

### Clinical and Research Implications

GLP-1RAs, particularly semaglutide, warrant further trials targeting alcohol consumption, cigarette consumption and cessation, and post-cessation weight gain. Evidence indicates therapeutic potential but does not establish their role in addiction treatment. Potential benefits for cardiometabolic and liver conditions warrant investigation, where GLP-1RAs could have pleiotropic benefits to reverse disease progression through alcohol consumption reduction^60^.

Future trials should distinguish treatment-seeking populations; assess consumption, craving, dependence and sustained abstinence over longer follow-up; report absolute effects; and use biochemical verification where feasible. Direct comparisons between GLP-1RA agents alongside established treatments are needed.

## CONCLUSIONS

Pooled estimates suggest that GLP-1RAs may reduce alcohol consumption and cigarettes smoked per day, with the clearest evidence for semaglutide. Effects on smoking cessation and clinical risk remain uncertain. Larger trials are needed to establish clinically meaningful benefit and sustained abstinence.

## Data Availability

All data produced in the present study are available upon reasonable request to the authors

---

**Figure 1.**
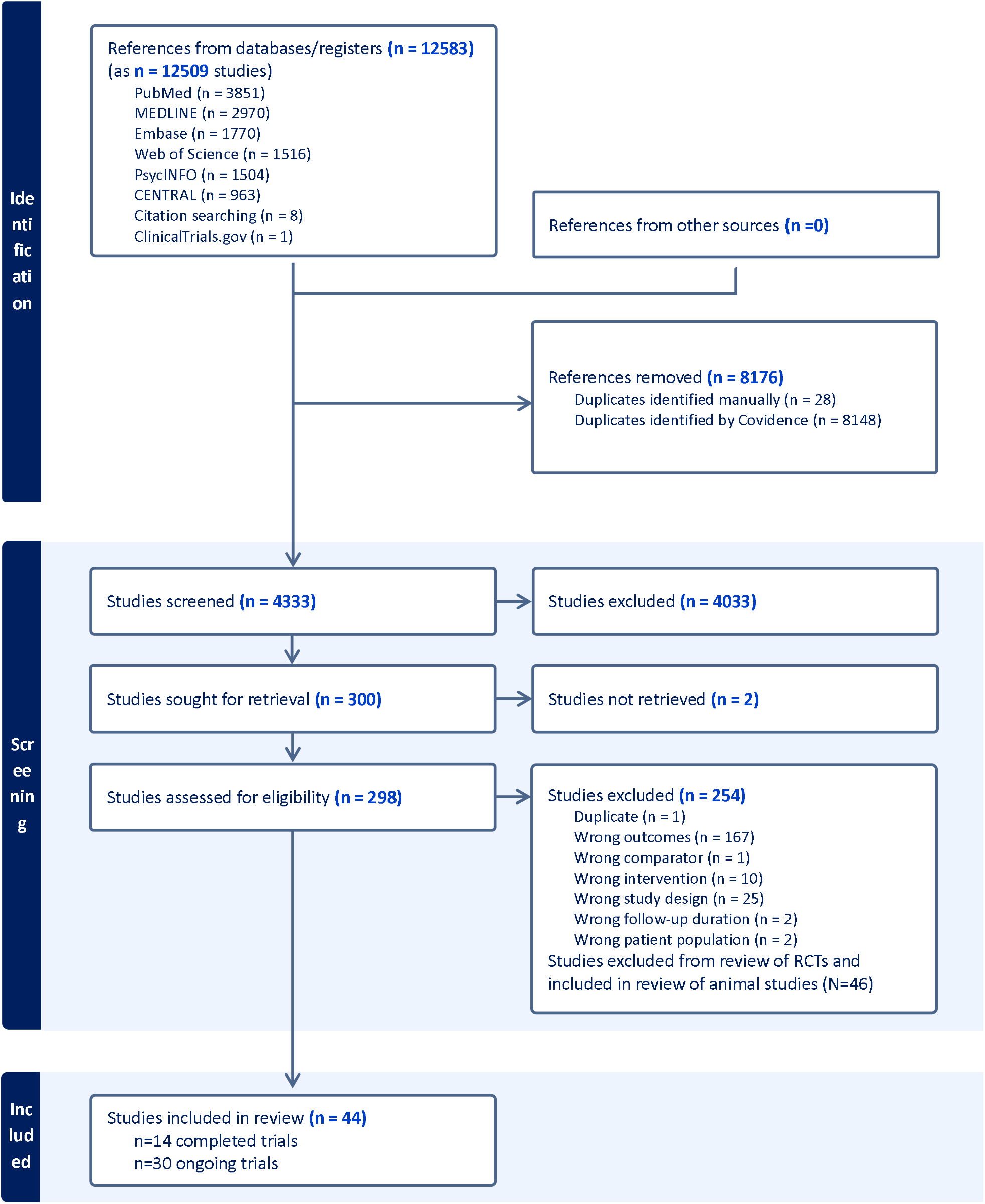
PRISMA flow diagram.

**Table 1.**
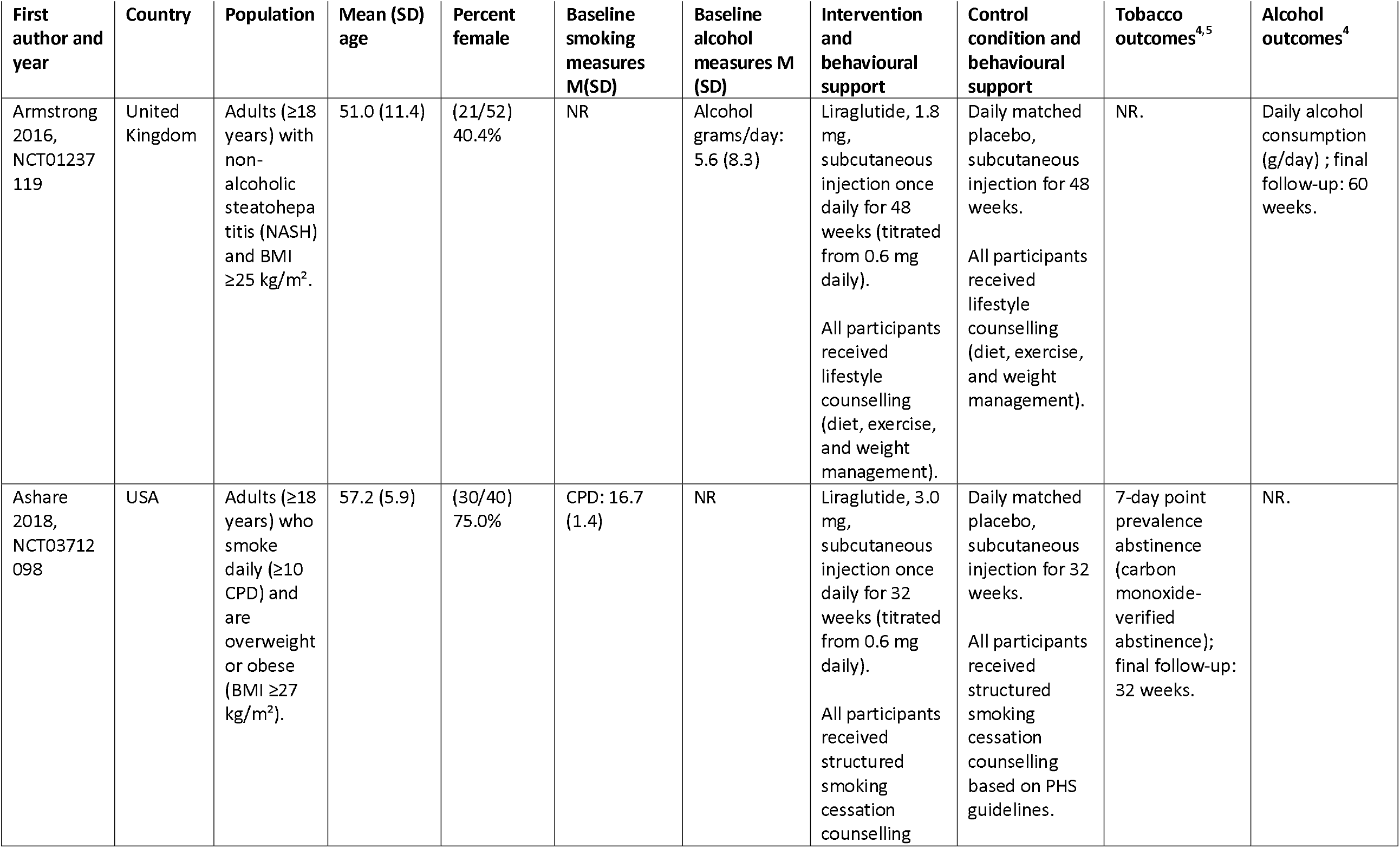
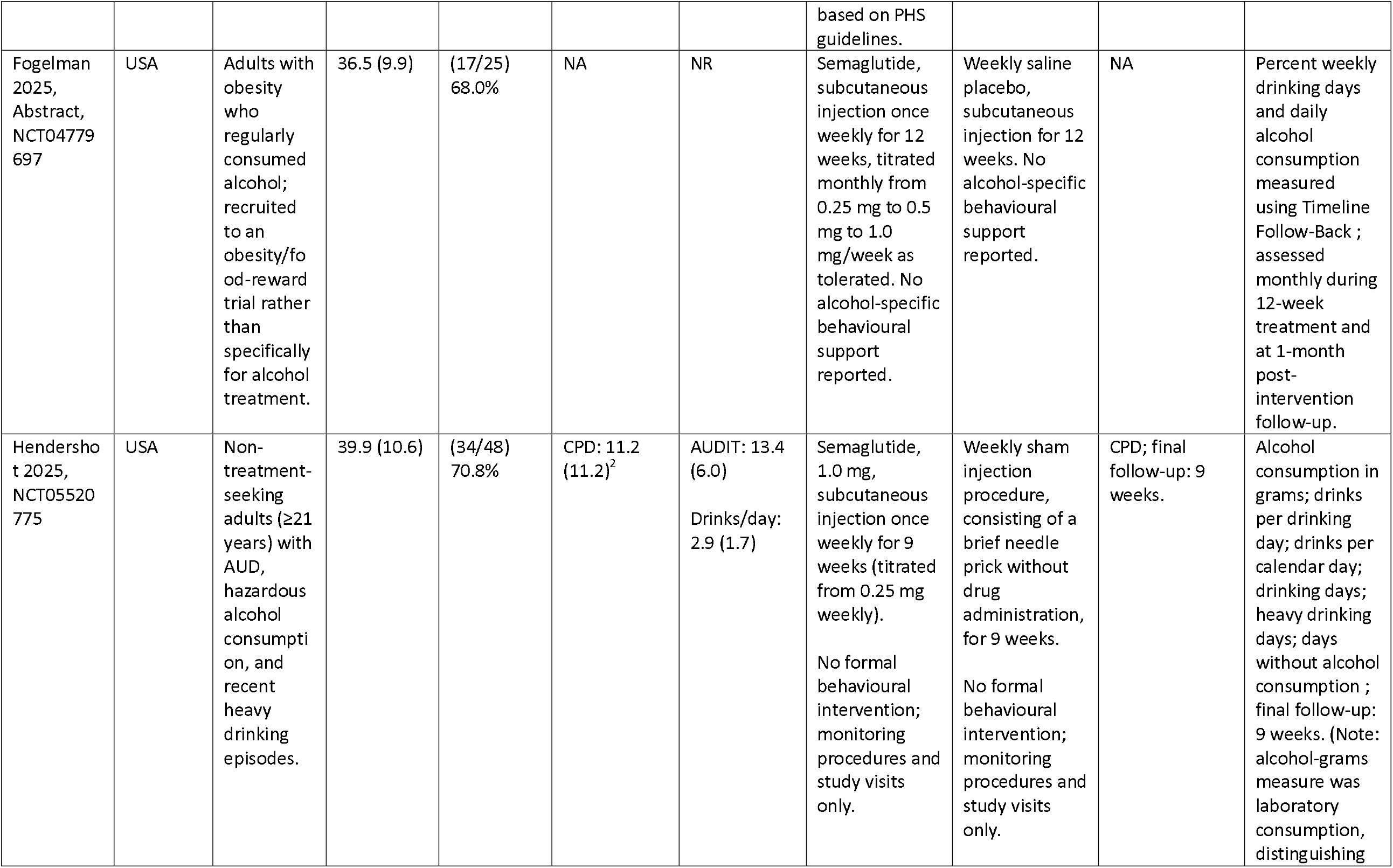
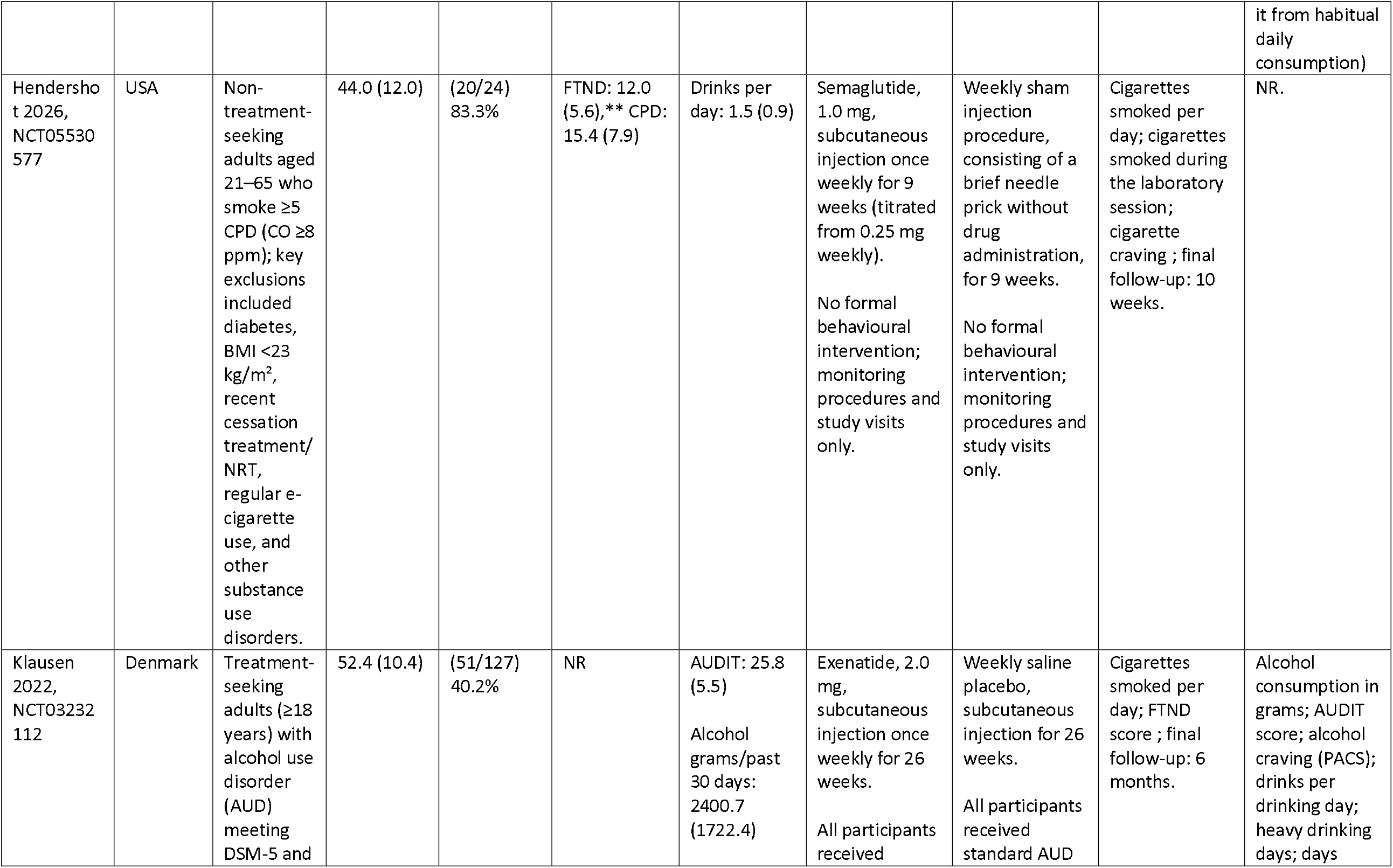
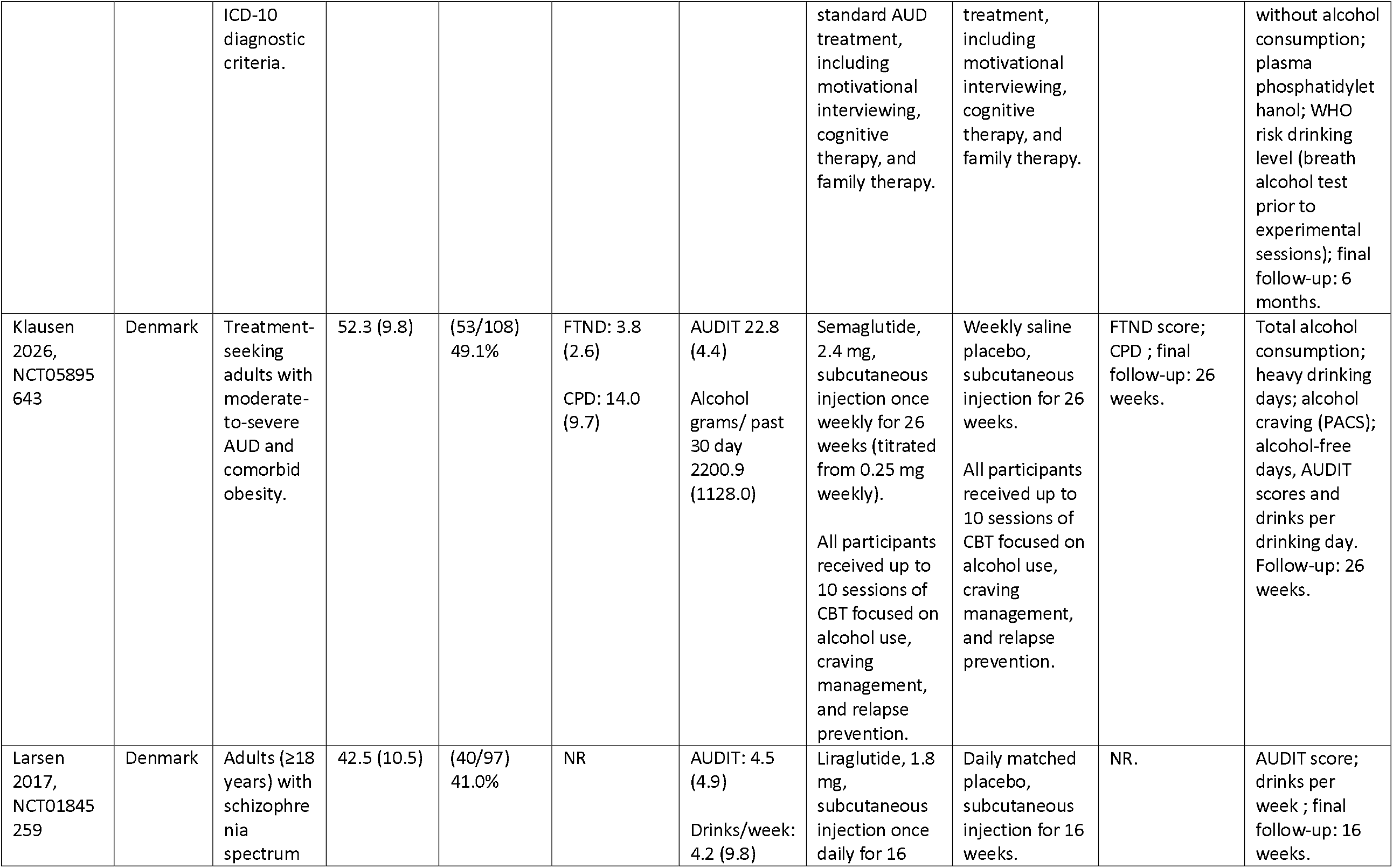
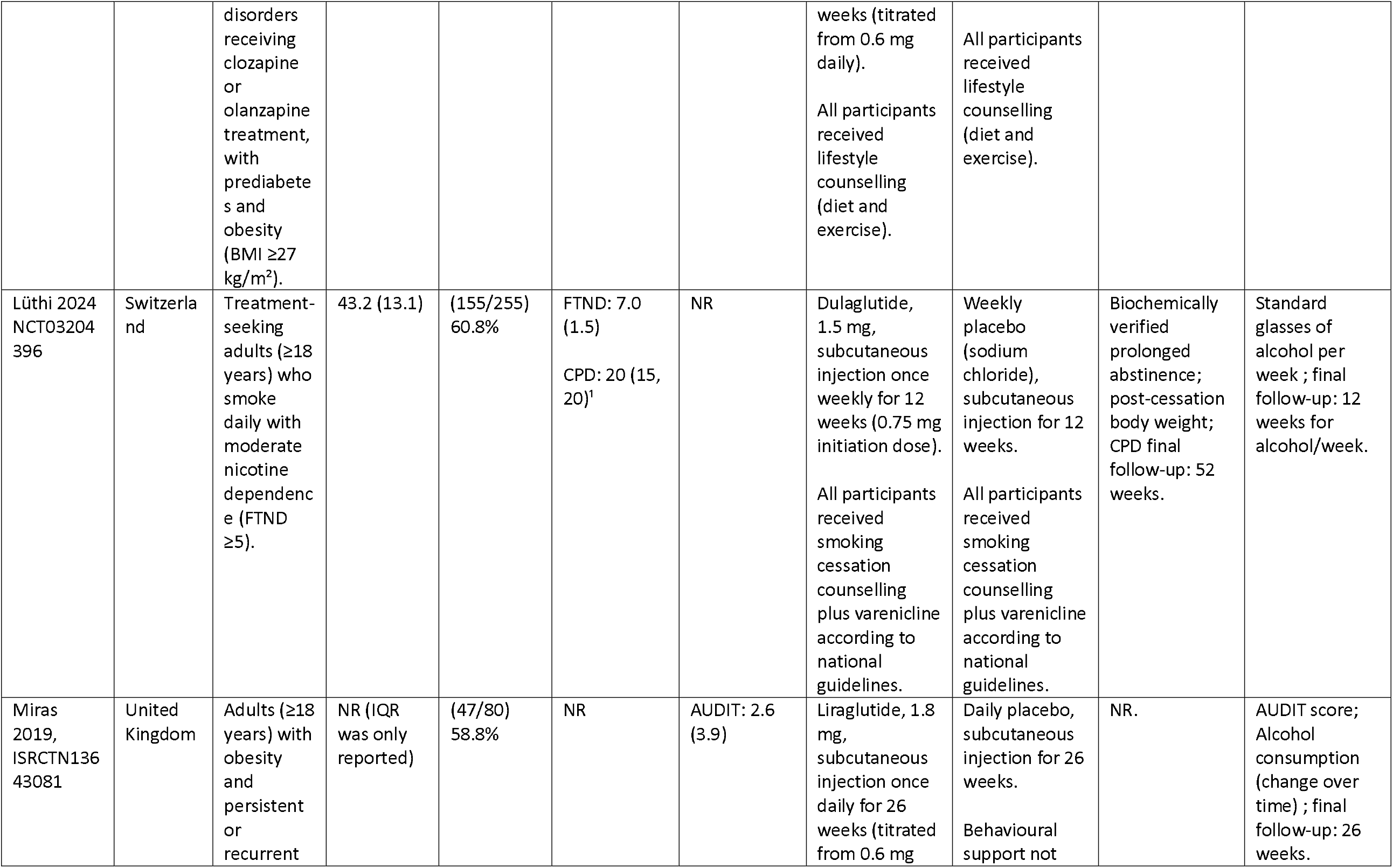
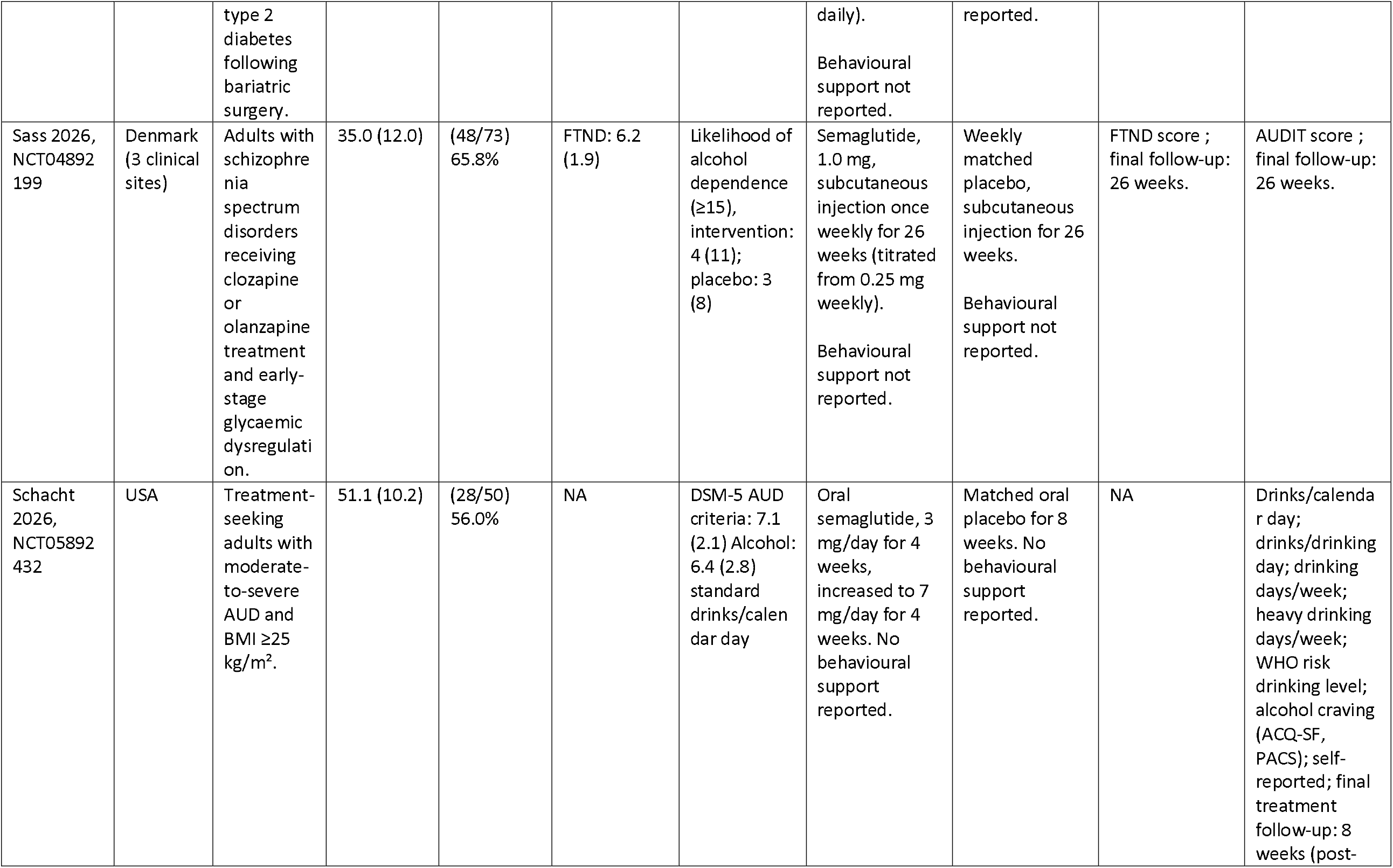
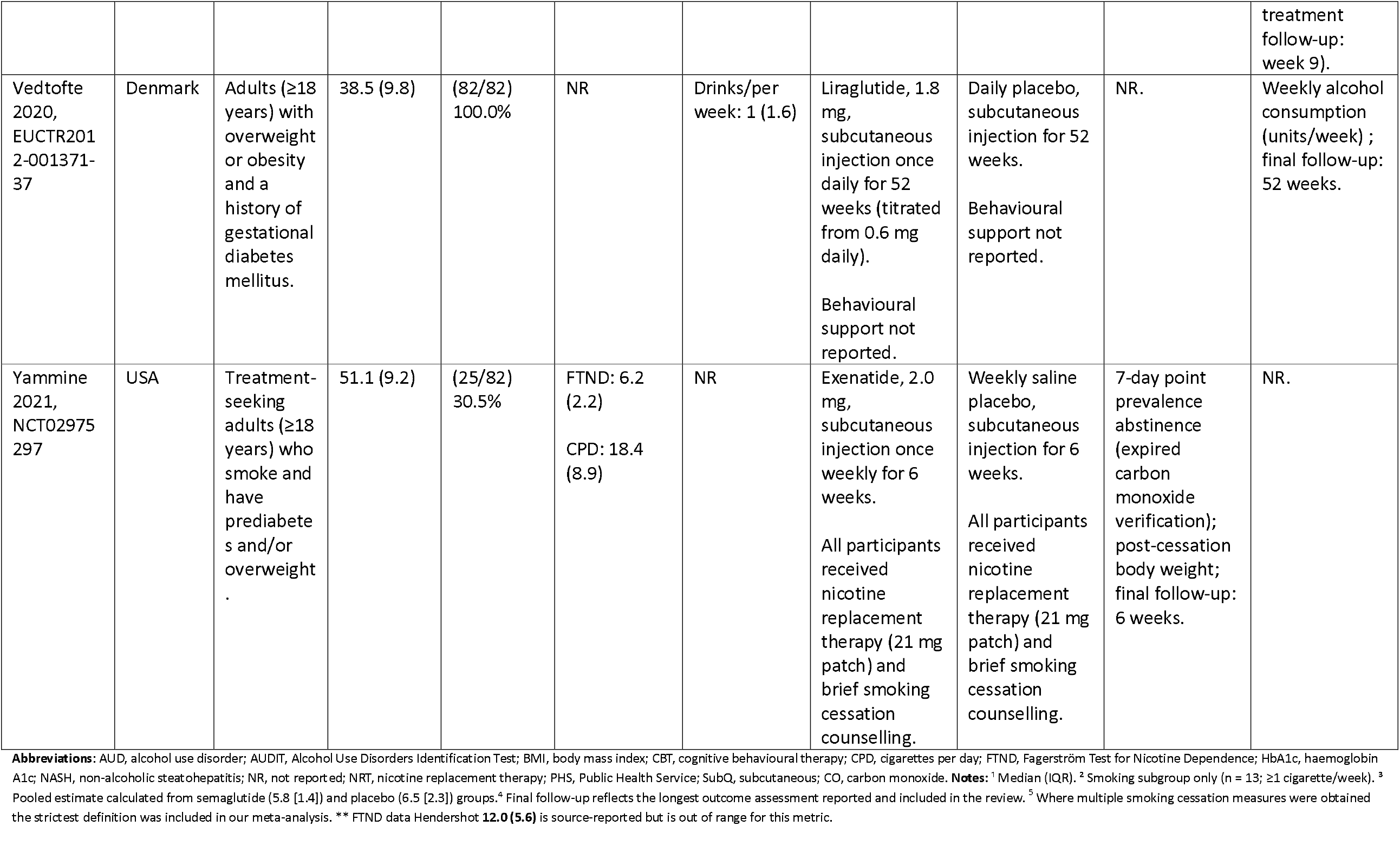
Study and participant characteristics for RCTs evaluating the effects of GLP-1RAs on smoking and alcohol related outcomes.

| First author and year | Country | Population | Mean (SD) age | Percent female | Baseline smoking measures M(SD) | Baseline alcohol measures M (SD) | Intervention and behavioural support | Control condition and behavioural support | Tobacco outcomes<sup>4,5</sup> | Alcohol outcomes<sup>4</sup> |
| --- | --- | --- | --- | --- | --- | --- | --- | --- | --- | --- |
| Armstrong 2016, NCT01237119 | United Kingdom | Adults (≥18 years) with non-alcoholic steatohepatitis (NASH) and BMI ≥25 kg/m<sup>2</sup>. | 51.0 (11.4) | (21/52) 40.4% | NR | Alcohol grams/day: 5.6 (8.3) | Liraglutide, 1.8 mg, subcutaneous injection once daily for 48 weeks (titrated from 0.6 mg daily).<br>All participants received lifestyle counselling (diet, exercise, and weight management). | Daily matched placebo, subcutaneous injection for 48 weeks.<br>All participants received lifestyle counselling (diet, exercise, and weight management). | NR. | Daily alcohol consumption (g/day) ; final follow-up: 60 weeks. |
| Ashare 2018, NCT03712098 | USA | Adults (≥18 years) who smoke daily (≥10 CPD) and are overweight or obese (BMI ≥27 kg/m<sup>2</sup>). | 57.2 (5.9) | (30/40) 75.0% | CPD: 16.7 (1.4) | NR | Liraglutide, 3.0 mg, subcutaneous injection once daily for 32 weeks (titrated from 0.6 mg daily).<br>All participants received structured smoking cessation counselling based on PHS guidelines. | Daily matched placebo, subcutaneous injection for 32 weeks.<br>All participants received structured smoking cessation counselling based on PHS guidelines. | 7-day point prevalence abstinence (carbon monoxide-verified abstinence); final follow-up: 32 weeks. | NR. |
| Fogelman 2025, Abstract, NCT04779697 | USA | Adults with obesity who regularly consumed alcohol; recruited to an obesity/food-reward trial rather than specifically for alcohol treatment. | 36.5 (9.9) | (17/25)<br>68.0% | NA | NR | Semaglutide, subcutaneous injection once weekly for 12 weeks, titrated monthly from 0.25 mg to 0.5 mg to 1.0 mg/week as tolerated. No alcohol-specific behavioural support reported. | Weekly saline placebo, subcutaneous injection for 12 weeks. No alcohol-specific behavioural support reported. | NA | Percent weekly drinking days and daily alcohol consumption measured using Timeline Follow-Back ; assessed monthly during 12-week treatment and at 1-month post-intervention follow-up. |
| Hendershot 2025, NCT05520775 | USA | Non-treatment-seeking adults (≥21 years) with AUD, hazardous alcohol consumption, and recent heavy drinking episodes. | 39.9 (10.6) | (34/48)<br>70.8% | CPD: 11.2 (11.2)<sup>2</sup> | AUDIT: 13.4 (6.0)<br>Drinks/day: 2.9 (1.7) | Semaglutide, 1.0 mg, subcutaneous injection once weekly for 9 weeks (titrated from 0.25 mg weekly).<br>No formal behavioural intervention; monitoring procedures and study visits only. | Weekly sham injection procedure, consisting of a brief needle prick without drug administration, for 9 weeks.<br>No formal behavioural intervention; monitoring procedures and study visits only. | CPD; final follow-up: 9 weeks. | Alcohol consumption in grams; drinks per drinking day; drinks per calendar day; drinking days; heavy drinking days; days without alcohol consumption ; final follow-up: 9 weeks. (Note: alcohol-grams measure was laboratory consumption, distinguishing it from habitual daily consumption) |
| Hendershot 2026, NCT05530577 | USA | Non-treatment-seeking adults aged 21–65 who smoke ≥5 CPD (CO ≥8 ppm); key exclusions included diabetes, BMI <23 kg/m², recent cessation treatment/ NRT, regular e-cigarette use, and other substance use disorders. | 44.0 (12.0) | (20/24) 83.3% | FTND: 12.0 (5.6), ** CPD: 15.4 (7.9) | Drinks per day: 1.5 (0.9) | Semaglutide, 1.0 mg, subcutaneous injection once weekly for 9 weeks (titrated from 0.25 mg weekly).<br>No formal behavioural intervention; monitoring procedures and study visits only. | Weekly sham injection procedure, consisting of a brief needle prick without drug administration, for 9 weeks.<br>No formal behavioural intervention; monitoring procedures and study visits only. | Cigarettes smoked per day; cigarettes smoked during the laboratory session; cigarette craving ; final follow-up: 10 weeks. | NR. |
| Klausen 2022, NCT03232112 | Denmark | Treatment-seeking adults (≥18 years) with alcohol use disorder (AUD) meeting DSM-5 and | 52.4 (10.4) | (51/127) 40.2% | NR | AUDIT: 25.8 (5.5)<br>Alcohol grams/past 30 days: 2400.7 (1722.4) | Exenatide, 2.0 mg, subcutaneous injection once weekly for 26 weeks.<br>All participants received | Weekly saline placebo, subcutaneous injection for 26 weeks.<br>All participants received standard AUD | Cigarettes smoked per day; FTND score ; final follow-up: 6 months. | Alcohol consumption in grams; AUDIT score; alcohol craving (PACS); drinks per drinking day; heavy drinking days; days |
|  |  | ICD-10 diagnostic criteria. |  |  |  |  | standard AUD treatment, including motivational interviewing, cognitive therapy, and family therapy. | treatment, including motivational interviewing, cognitive therapy, and family therapy. |  | without alcohol consumption; plasma phosphatidylethanol; WHO risk drinking level (breath alcohol test prior to experimental sessions); final follow-up: 6 months. |
| Klausen 2026, NCT05895643 | Denmark | Treatment-seeking adults with moderate-to-severe AUD and comorbid obesity. | 52.3 (9.8) | (53/108)<br>49.1% | FTND: 3.8 (2.6)<br>CPD: 14.0 (9.7) | AUDIT 22.8 (4.4)<br>Alcohol grams/ past 30 day 2200.9 (1128.0) | Semaglutide, 2.4 mg, subcutaneous injection once weekly for 26 weeks (titrated from 0.25 mg weekly).<br>All participants received up to 10 sessions of CBT focused on alcohol use, craving management, and relapse prevention. | Weekly saline placebo, subcutaneous injection for 26 weeks.<br>All participants received up to 10 sessions of CBT focused on alcohol use, craving management, and relapse prevention. | FTND score; CPD ; final follow-up: 26 weeks. | Total alcohol consumption; heavy drinking days; alcohol craving (PACS); alcohol-free days, AUDIT scores and drinks per drinking day. Follow-up: 26 weeks. |
| Larsen 2017, NCT01845259 | Denmark | Adults (≥18 years) with schizophrenia spectrum | 42.5 (10.5) | (40/97)<br>41.0% | NR | AUDIT: 4.5 (4.9)<br>Drinks/week: 4.2 (9.8) | Liraglutide, 1.8 mg, subcutaneous injection once daily for 16 | Daily matched placebo, subcutaneous injection for 16 weeks. | NR. | AUDIT score; drinks per week ; final follow-up: 16 weeks. |
| | | disorders receiving clozapine or olanzapine treatment, with prediabetes and obesity (BMI $\geq 27$ kg/m<sup>2</sup>). | | | | | weeks (titrated from 0.6 mg daily).<br>All participants received lifestyle counselling (diet and exercise). | All participants received lifestyle counselling (diet and exercise). | | |
| Lüthi 2024<br>NCT03204396 | Switzerland | Treatment-seeking adults ($\geq 18$ years) who smoke daily with moderate nicotine dependence (FTND $\geq 5$). | 43.2 (13.1) | (155/255)<br>60.8% | FTND: 7.0 (1.5)<br>CPD: 20 (15, 20)<sup>1</sup> | NR | Dulaglutide, 1.5 mg, subcutaneous injection once weekly for 12 weeks (0.75 mg initiation dose).<br>All participants received smoking cessation counselling plus varenicline according to national guidelines. | Weekly placebo (sodium chloride), subcutaneous injection for 12 weeks.<br>All participants received smoking cessation counselling plus varenicline according to national guidelines. | Biochemically verified prolonged abstinence; post-cessation body weight; CPD final follow-up: 52 weeks. | Standard glasses of alcohol per week ; final follow-up: 12 weeks for alcohol/week. |
| Miras 2019,<br>ISRCTN13643081 | United Kingdom | Adults ($\geq 18$ years) with obesity and persistent or recurrent | NR (IQR was only reported) | (47/80)<br>58.8% | NR | AUDIT: 2.6 (3.9) | Liraglutide, 1.8 mg, subcutaneous injection once daily for 26 weeks (titrated from 0.6 mg | Daily placebo, subcutaneous injection for 26 weeks.<br>Behavioural support not | NR. | AUDIT score; Alcohol consumption (change over time) ; final follow-up: 26 weeks. |
|  |  | type 2 diabetes following bariatric surgery. |  |  |  |  | daily).<br>Behavioural support not reported. | reported. |  |  |
| Sass 2026, NCT04892 199 | Denmark (3 clinical sites) | Adults with schizophrenia spectrum disorders receiving clozapine or olanzapine treatment and early-stage glycaemic dysregulation. | 35.0 (12.0) | (48/73) 65.8% | FTND: 6.2 (1.9) | Likelihood of alcohol dependence ($\geq 15$), intervention: 4 (11); placebo: 3 (8) | Semaglutide, 1.0 mg, subcutaneous injection once weekly for 26 weeks (titrated from 0.25 mg weekly).<br>Behavioural support not reported. | Weekly matched placebo, subcutaneous injection for 26 weeks.<br>Behavioural support not reported. | FTND score ; final follow-up: 26 weeks. | AUDIT score ; final follow-up: 26 weeks. |
| Schacht 2026, NCT05892 432 | USA | Treatment-seeking adults with moderate-to-severe AUD and BMI $\geq 25$ kg/m<sup>2</sup>. | 51.1 (10.2) | (28/50) 56.0% | NA | DSM-5 AUD criteria: 7.1 (2.1) Alcohol: 6.4 (2.8) standard drinks/calendar day | Oral semaglutide, 3 mg/day for 4 weeks, increased to 7 mg/day for 4 weeks. No behavioural support reported. | Matched oral placebo for 8 weeks. No behavioural support reported. | NA | Drinks/calendar day; drinks/drinking day; drinking days/week; heavy drinking days/week; WHO risk drinking level; alcohol craving (ACQ-SF, PACS); self-reported; final treatment follow-up: 8 weeks (post- treatment follow-up: week 9). |
| Vedtofte 2020, EUCTR2012-001371-37 | Denmark | Adults (≥18 years) with overweight or obesity and a history of gestational diabetes mellitus. | 38.5 (9.8) | (82/82) 100.0% | NR | Drinks/per week: 1 (1.6) | Liraglutide, 1.8 mg, subcutaneous injection once daily for 52 weeks (titrated from 0.6 mg daily).<br>Behavioural support not reported. | Daily placebo, subcutaneous injection for 52 weeks.<br>Behavioural support not reported. | NR. | Weekly alcohol consumption (units/week) ; final follow-up: 52 weeks. |
| Yammine 2021, NCT02975297 | USA | Treatment-seeking adults (≥18 years) who smoke and have prediabetes and/or overweight. | 51.1 (9.2) | (25/82) 30.5% | FTND: 6.2 (2.2)<br>CPD: 18.4 (8.9) | NR | Exenatide, 2.0 mg, subcutaneous injection once weekly for 6 weeks.<br>All participants received nicotine replacement therapy (21 mg patch) and brief smoking cessation counselling. | Weekly saline placebo, subcutaneous injection for 6 weeks.<br>All participants received nicotine replacement therapy (21 mg patch) and brief smoking cessation counselling. | 7-day point prevalence abstinence (expired carbon monoxide verification); post-cessation body weight; final follow-up: 6 weeks. | NR. |

**Abbreviations:** AUD, alcohol use disorder; AUDIT, Alcohol Use Disorders Identification Test; BMI, body mass index; CBT, cognitive behavioural therapy; CPD, cigarettes per day; FTND, Fagerström Test for Nicotine Dependence; HbA1c, haemoglobin A1c; NASH, non-alcoholic steatohepatitis; NR, not reported; NRT, nicotine replacement therapy; PHS, Public Health Service; SubQ, subcutaneous; CO, carbon monoxide. **Notes:** <sup>1</sup> Median (IQR). <sup>2</sup> Smoking subgroup only (n = 13; ≥1 cigarette/week). <sup>3</sup> Pooled estimate calculated from semaglutide (5.8 [1.4]) and placebo (6.5 [2.3]) groups. <sup>4</sup> Final follow-up reflects the longest outcome assessment reported and included in the review. <sup>5</sup> Where multiple smoking cessation measures were obtained the strictest definition was included in our meta-analysis. \*\* FTND data Hendershot **12.0 (5.6)** is source-reported but is out of range for this metric.

**Table 2.**
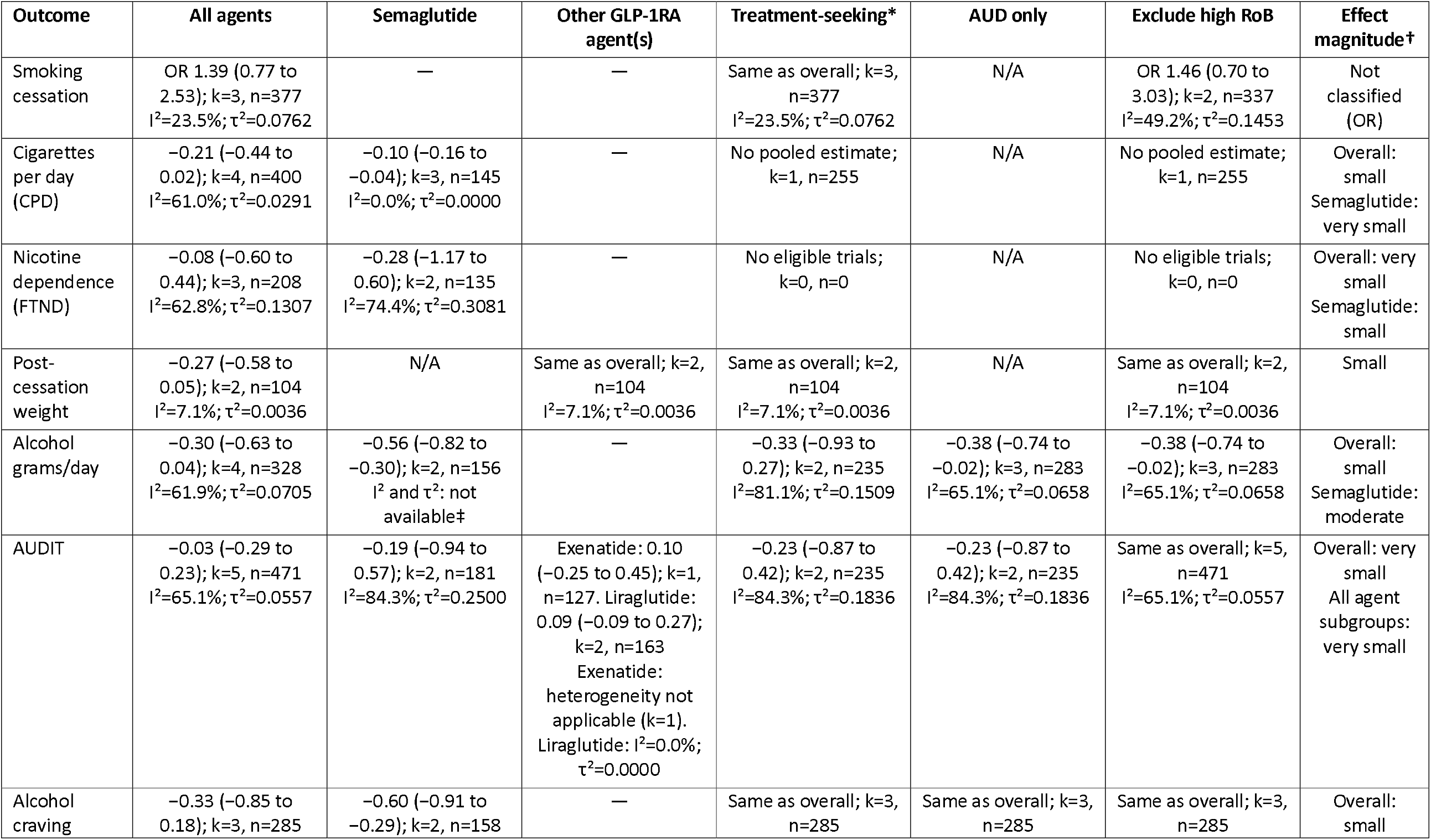
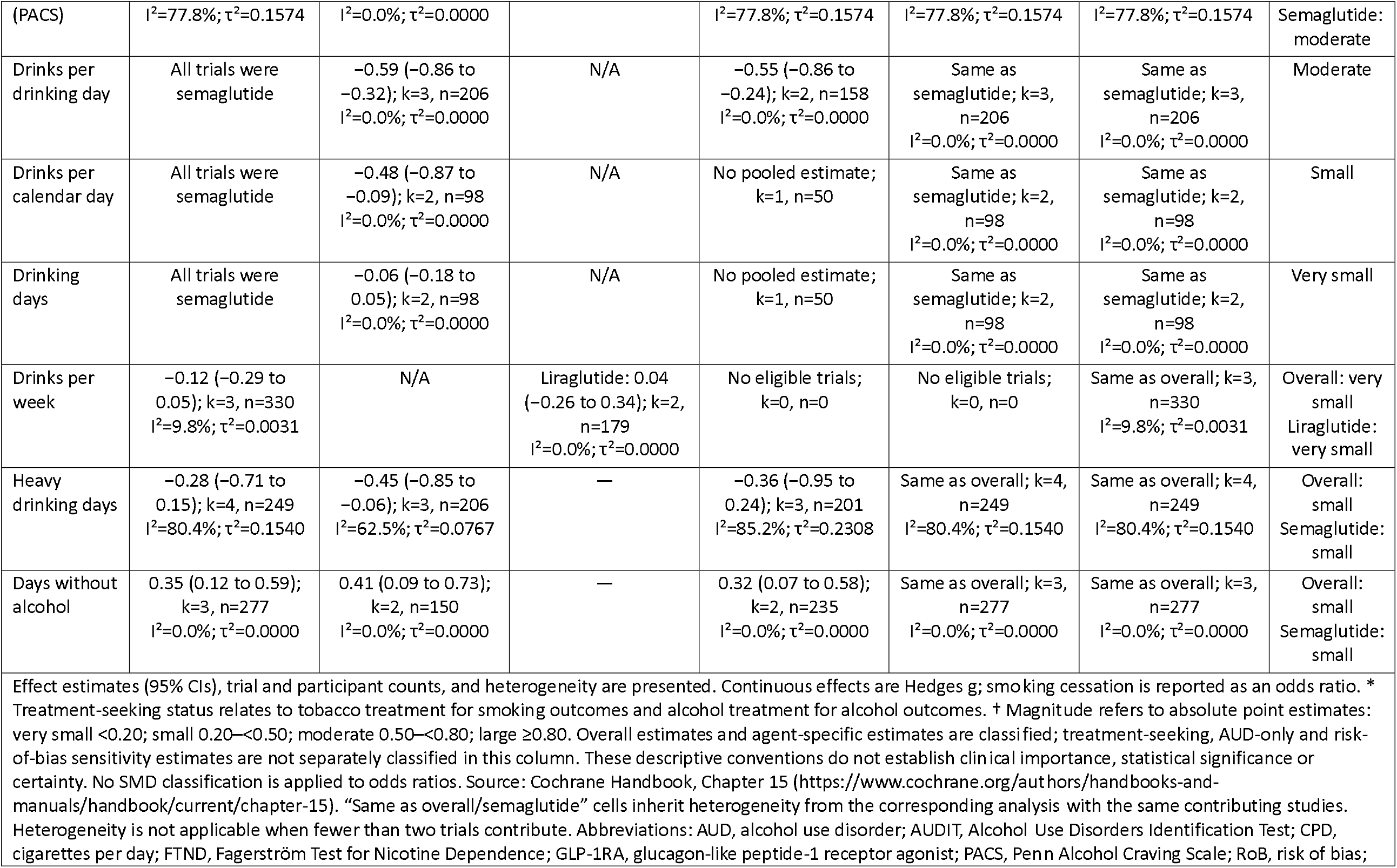
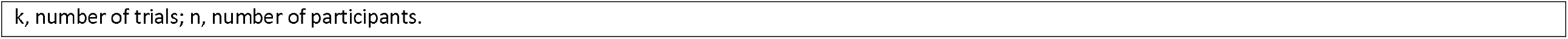
Pooled estimates for effects of GLP-1RAs on outcomes: effect estimates presented by sensitivity analysis.

**Figure 1.**
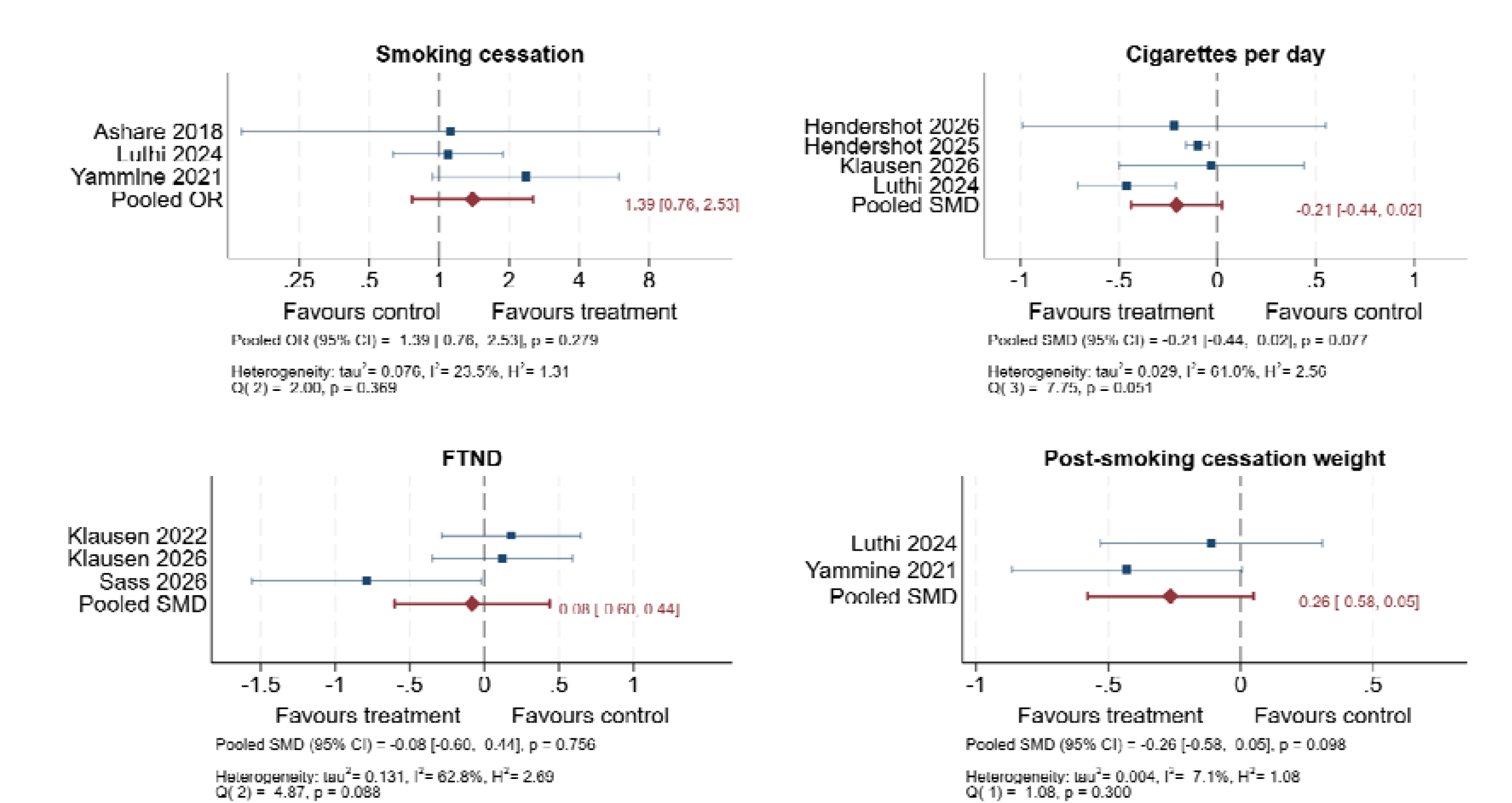
Forest plots showing the effects of any GLP-1RA on smoking-related outcomes*. *Plots show either pooled standardised mean difference (SMDs) or odds ratio (OR) depending on outcome. Panels retain outcome-specific direction of effect. FTND = Fagerström Test for Nicotine Dependence.

**Figure 1.**
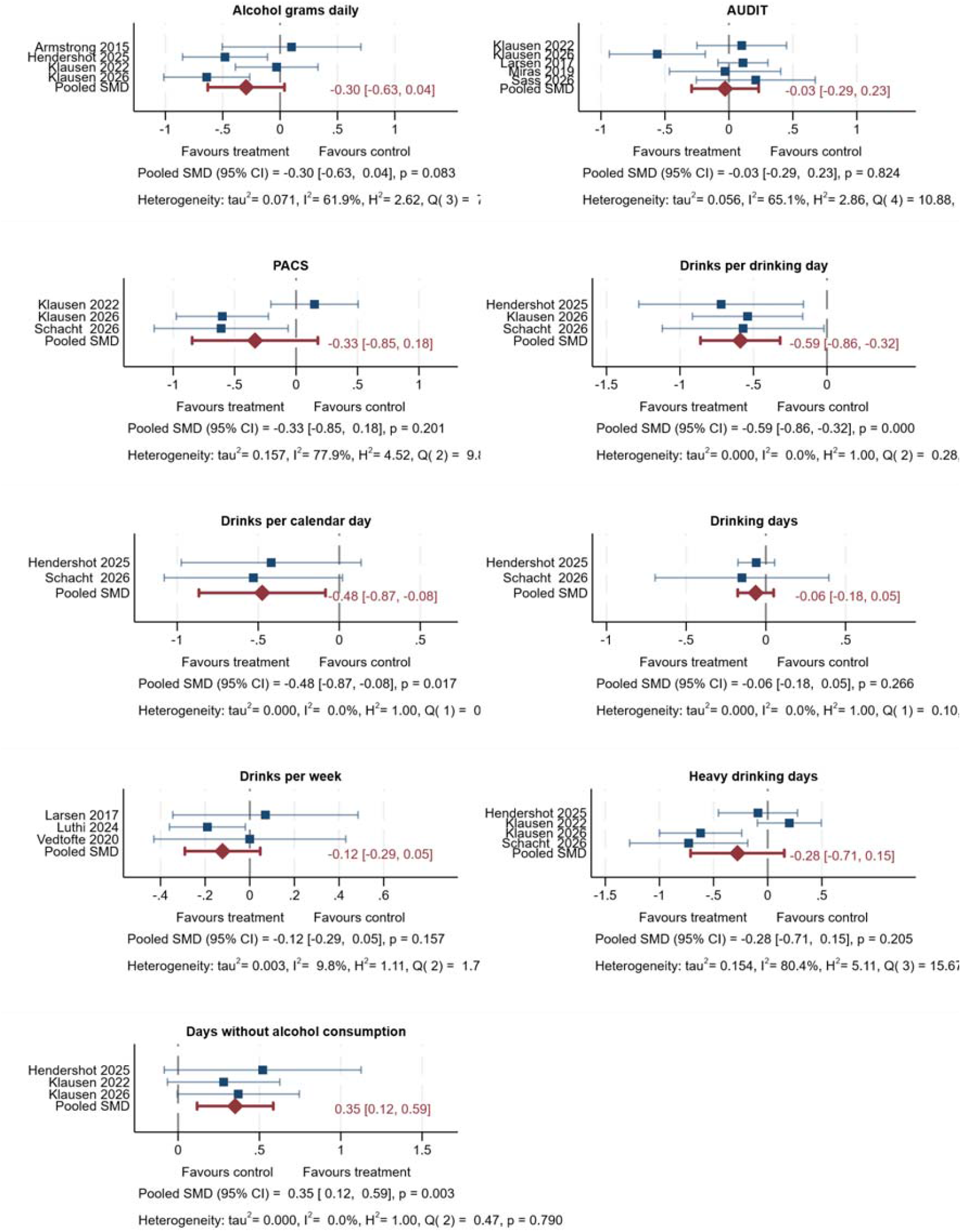
Forest plots showing the effects of any GLP-1RA on alcohol-related outcomes*. *Plots show the pooled standardised mean difference (SMDs). Panels retain outcome-specific direction of effect. AUDIT = Alcohol use disorders identification test. PACS =Penn Alcohol Craving Scale

## Notes

**FUNDING** This study was partly funded by the University of Bath Research Support fund. GT is funded by an NIHR Postdoctoral Fellowship. GT acknowledges additional funding from CRUK and the Causality in Healthcare AI Hub, funded by EPSRC and UKRI, and reports funding from the NIHR Bristol Biomedical Research Centre, University Hospitals Bristol and Weston NHS Foundation Trust, and the University of Bristol (NIHR203315). BC acknowledges a Mary and Peter Childs doctoral research support award. DT is funded as an NIHR Academic Clinical Lecturer within the Centre for Academic Mental Health at the University of Bristol. CB is supported by an NIHR-funded postdoctoral position within the NIHR Mental Health Research Group at the University of Bath (reference NIHR207583). IF is supported by a BBSRC-DTP studentship (DTP3 2020-2025 entry - BB/T008725/1). SJ is supported by an international student scholarship from the University of Bath. MH supported by NIHR HPRU Evaluation and Behavioural Science.

### Competing Interest Statement

GT worked at a health economics and outcomes research company from 2022 to 2024, and their clients were pharmaceutical companies. Some of these clients were makers of GLP-1RA medicines. GT has no relationships with these companies and they were not involved in this project. KA has received consultancy fees from Novo Nordisk and lecture honoraria from Madrigal and Norgine.

### Summary of Updates

I added semi-colons to the abstract section on the medRxiv online form to improve readability, i.e., ":". Added an effect size interpretation column to Table 2. Updated Matthew Hickman's affiliations.

