## Supplementary material for "GLP-1 Receptor Agonists (GLP-1RAs) for treatment of tobacco and alcohol dependence: a systematic review and meta-analysis of randomised controlled trials": e-Supplement

### LIST OF TABLES AND FIGURES

#### TABLES

**eTable 9 Exploratory tests for small-study effects across smoking and alcohol outcomes** .. 39

#### FIGURES

#### *1. METHODS*

##### 1.1. Eligibility Criteria: Population, Interventions, Comparators, and Outcomes (PICO)

**eTable 1 Eligibility criteria**

| PICO element | Criteria |
|---|---|
| Population | People of any age who smoke tobacco or consume alcohol, including those with comorbidities such as type 2 diabetes or people who are overweight. |
| Intervention | Any GLP-1RA, GIP RA, or dual GLP-1RA/GIP RA, at any dose, route or duration. |
| Comparator | Placebo, no treatment, usual care, or another active intervention, as applicable. |
| Outcomes | Smoking related: Smoking cessation (any definition) cigarettes smoked per day, nicotine dependence, post-cessation weight.<br><br>Alcohol related: Alcohol consumption, alcohol abstinence, alcohol craving and alcohol risk (e.g., AUDIT) |
| Follow-up | At least 28 days. For each outcome, we selected the longest available follow-up of at least 28 days for which sufficient data were available for analysis. This could precede the trial's final follow-up. |
| Study design and eligibility | RCTs in any language or setting. Abstracts, registry results, and unpublished studies were included when usable data were available. |

##### 1.2. Search Strategy

#1 GLP-1 receptor agonist terms: ("glucagon-like peptide 1" OR GLP-1 OR GLP1 OR "glucagon-like peptide-1 receptor agonist*" OR liraglutide OR semaglutide OR exenatide OR dulaglutide OR lixisenatide OR albiglutide OR efpeglenatide)

#2 Addiction terms: (smoking OR "tobacco use" OR "nicotine dependence" OR nicotine OR "smoking cessation" OR "tobacco cessation" OR "alcohol use" OR "alcohol drinking" OR "alcohol consumption" OR "alcohol dependence" OR "alcohol use disorder*" OR AUD OR ethanol)

#3 Study Type – Human RCTs: (randomised controlled trial[Publication Type] OR randomised[Title/Abstract] OR placebo[Title/Abstract] OR clinical trial [Title/Abstract]) OR control*

#4 Study Type – Animal Studies: ("animal experimentation"[MeSH Terms] OR "animal model*" OR mice OR mouse OR rat OR rats OR rodent* OR "preclinical study" OR "non-human primate*" OR monkey OR monkeys)

#1 AND #2 AND (#3 OR #4)

##### 1.3. Measures of Treatment Effect

Continuous outcomes were expressed as Hedges *g* standardised mean differences with 95%CIs, and dichotomous outcomes as odds ratios with 95%CIs.

Formula 1:
$$J \approx 1 - \frac{3}{4(n_1 + n_2) - 9}$$

$n_1$ and $n_2$ are the sample sizes of Group 1 and Group 2.

Formula 2:
$$\text{Hedges}'\ g = J \times \text{SMD}$$

Formula3:
$$SE_g = SE_{SMD} \times J$$

$$\text{Lower Limit (LL)} = g - (1.96 \times SE_g)$$

Formula 4:
$$\text{Upper Limit (UL)} = g + (1.96 \times SE_g)$$

##### 1.4. Data Synthesis

We conducted separate meta-analyses for clinically comparable smoking and alcohol outcomes. We pooled data from similar interventions and outcomes.

For each study, for any outcome measured using continuous data, we calculated the SMD and 95%CIs in change in outcomes from baseline to follow-up, between those in the intervention and control arms. We calculated the SMD (95%CI) by extracting any of the following data, in order of preference:

1. Adjusted or unadjusted mean difference (MD; difference in change from baseline to follow-up) and measure of variation between treatment groups, with a preference for adjusted estimates;
2. Mean change in outcome scores from baseline to follow-up and measure of variance, by treatment group;
3. Mean outcome scores and measures of variance at baseline and final follow-up, by treatment group.

Where type (3) data were collected we then calculated the mean change and its variance for each exposure group[1]; for data types (2) and (3) we calculated the SMD, using standard formulae outlined within the Cochrane Handbook for Systematic Reviews of Interventions[2]. We assumed a within-participant correlation of 0.59 between baseline and post-treatment scores, based on the median reported across research areas in the NICE Guideline Methodology Guidelines[3] and consistent with previous reviews conducted by our group[4,5]. We sought statistical support where appropriate.

For each study, for any outcome measured using dichotomous data, we calculated ORs and the 95%CI. We calculated the ORs (95%CI) by extracting any of the following data, in order of preference:

1. Adjusted or unadjusted ORs and a measure of variance;
2. Other types of effect estimate that could be converted into ORs (i.e. risk ratios, hazard ratios);
3. We extracted data to calculate ORs for outcomes and the variance (N participants intervention group at follow-up, N participants with outcome in control group at follow-up, N participants in quit group at follow-up, N participants with outcome in quit group at follow up).

We then logged ORs using a standard formula before inputting them into meta-analyses[2]. We sought statistical support to help with the above calculations where appropriate.

For continuous outcomes, we pooled SMDs (95%CI) across individual studies using a generic inverse variance random-effects model. An SMD of greater than 0 indicated that outcome was greater in the intervention group, whereas an SMD of less than 0 indicated the outcome was greater in the control group. For dichotomous outcomes, we pooled ORs (95%CI) across individual studies using a random-effects model. An OR of greater than 1 indicated the outcome was more frequent in the intervention group at follow-up, and less than 1 indicated that the outcome was more frequent in the control group. We conducted meta-analyses of the SMD and OR for each outcome separately using Stata package meta[6].

##### 1.5. Assessment of Heterogeneity and Reporting Biases

Statistical heterogeneity was assessed using the $I^2$ and $Chi^2$ statistics and visually through forest plots. Sources of heterogeneity were explored via subgroup and sensitivity analyses where there were sufficient studies to do so.

For each meta-analysed outcome, we generated a funnel plot and, where at least 3 studies were available, calculated a two-sided Egger regression test. Cochrane recommends testing for funnel-plot asymmetry only when at least 10 studies contribute to a meta-analysis, because tests have low power with fewer studies[2]. As all our analyses contained fewer than 10 studies, we present the plots and tests for illustration only and draw no conclusions about small-study effects or publication bias from them[2]. We did not run Egger's test for outcomes with only 2 studies. Conventional Egger regression also requires caution here because odds ratios and SMDs can be correlated with their standard errors.

##### 1.6. Dealing with Missing Data

We contacted study authors for missing or unclear outcome data and report the results of these requests in the Appendix.

##### 1.7. Protocol Deviations

The pre-registered protocol covered both human RCTs and animal experiments[7]; this paper reports the RCTs, and the animal evidence will be reported separately. Where a trial reported an outcome at multiple time points, we used the longest follow-up for which that outcome and sufficient data for analysis were available. Consequently, the time point included in a meta-analysis could be earlier than the trial's final follow-up. We additionally synthesised nicotine dependence, cigarettes per day (CPD) and alcohol craving where trial data permitted. Prespecified secondary outcomes without sufficient usable data were not pooled. Indirect and population-adjusted comparisons were not applicable because the included trials did not provide suitable active-comparator evidence. Planned subgroup and sensitivity analyses were limited by the number of trials per outcome; the analyses performed are reported in the Results section.

#### 2. RESULTS

##### 2.1 Study Characteristics

We included 14 parallel, 2-arm, placebo-controlled RCTs (**eTable 2**). Populations were heterogeneous, including people with metabolic conditions, treatment-seeking and non-treatment-seeking smokers, people with alcohol use disorder, and those with schizophrenia spectrum disorders. Trials evaluated liraglutide, semaglutide, exenatide or dulaglutide over 6-52 weeks, with substantial variation in dose, route of administration and behavioural support; industry funding or related conflicts of interest were common. Eight trials reported smoking outcomes, 11 reported alcohol outcomes and 5 reported both, with 13 trials comprising 1124 participants contributing to at least one meta-analysis. See **eTable *3*** for additional study characteristics. A further 30 ongoing studies or completed studies awaiting results indicate that this remains a rapidly developing evidence base (**eTable 4**). For a list of excluded studies see **eTable 5**.

##### 2.2 Included Studies

Studies were conducted across a range of high-income settings in Europe and North America. Five studies were undertaken in Denmark[8-12], 6 in the United States[13-18], 2 in the United Kingdom[19,20], and 1 in Switzerland[21]. Six trials received industry funding or support [9-11,13,19,20], and 11 of the 12 trials with available disclosure information reported at least one industry-related author conflict of interest[8-12,15,16,18-21]; Yammine et al. (2021)[17] declared no conflicts of interest, while COI information was unavailable for Ashare[13] and Fogelman et al (2025)[14].

Data extracted were predominantly published in peer-reviewed journals, except for 1 trial where data were obtained from open access trial registry[13], and 1 conference[14]. Study authors were contacted to obtain or clarify unpublished outcome data required for meta-analysis; additional data were obtained for Hendershot et al[15] and Lüthi et al[21], whereas requests relating to Schacht et al[18], Fogelman et al[14] and Sarma et al[22] did not yield usable unpublished data (Although Schacht[18] was published after our initial request). For further details see **eTable 6**.

##### 2.3 Participants

Human RCTs: This review includes 1149 human participants over 14 trials, with 1124 included in the meta-analyses; this excludes Fogelman et al. (2025; n=25), which was included only in the narrative synthesis because sufficient numerical outcome data were unavailable.

Across the 14 human trials, 10 included participants with metabolic conditions or elevated metabolic risk, including NASH, obesity, diabetes, prediabetes or previous gestational diabetes[9-14,17-20]. Four trials specifically recruited daily smokers. Three recruited treatment-seeking smokers[13,17,21], whereas 1 recruited smokers who were not seeking cessation treatment[16]. One trial recruited non-treatment-seeking participants with AUD and assessed smoking among participants who smoked[15], rather than recruiting on the basis of smoking status. Four trials specifically recruited participants with AUD: three recruited treatment-seeking participants[8,12,18] whereas 1 recruited non-treatment-seeking participants[15]. Some trials excluded participants receiving addiction treatment or with comorbid substance use disorders[8,13,15,17,23].

Mean participant age ranged from 35.0 years[10] to 57.2 years[13], with the majority of studies recruiting predominantly middle-aged samples. Female representation ranged from 30.5%[17] to 100% in the women-only study by Vedtofte et al. (2020)[11]. Six studies reported alcohol outcomes only[9,11,14,18-20], 3 reported tobacco outcomes only[13,16,17], and 5 reported both alcohol and tobacco outcomes[8,10,12,15,21]. Among studies reporting valid Fagerström Test for Nicotine Dependence (FTND) scores, baseline means ranged from 3.8 (SD 2.6) to 7.0 (SD 1.5)[12,21]. Baseline cigarette consumption ranged from a mean of 11.2 cigarettes per day (CPD) to a median of 20 CPD[15,21]. Baseline Alcohol Use Disorders Identification Test (AUDIT) scores ranged from 2.6 (SD 3.9) to 25.8 (SD 5.5)[8,20]. Alcohol consumption could not be expressed as a single meaningful range because studies reported different measures and timeframes, including drinks per day[15,18], drinks per week[9,11], grams per day[19] and grams over the preceding 30 days[8,12].

##### 2.4 Intervention

Across the 14 included trials, interventions varied by GLP-1RA agent, dose, duration, and level of behavioural support. Five studies evaluated liraglutide[9,11,13,19,20], 6 evaluated semaglutide[10,12,14-16,18], 2 evaluated exenatide[8,17], and 1 evaluated dulaglutide[21]. All interventions except that of Schacht et al[18] were delivered by subcutaneous injection. Liraglutide was administered subcutaneously once daily, titrated from 0.6 mg to 1.8 or 3.0 mg, for 16-52 weeks. Injectable semaglutide was administered once weekly, titrated from 0.25 mg to 1.0 or 2.4 mg, for 9-26 weeks; oral semaglutide was administered daily for 8 weeks, increasing from 3 mg to 7 mg after 4 weeks. Exenatide was administered subcutaneously once weekly at 2.0 mg for 6 or 26 weeks, while dulaglutide was administered once weekly for 12 weeks, increasing from 0.75 to 1.5 mg.

Where behavioural support was provided, the same level and type of support was offered to participants in both the intervention and control groups. Two studies provided no formal addiction-focused behavioural treatment beyond monitoring, questionnaires and study visits[15,16]. Three smoking-cessation studies provided structured behavioural counselling[13,17,21], with 2 also providing cessation pharmacotherapy: varenicline[21] and NRT[17]. Two alcohol-focused studies incorporated standard alcohol-use disorder treatment: motivational interviewing, cognitive therapy and family therapy[8], and structured cognitive behavioural therapy addressing motivation, craving, coping strategies and relapse prevention[12]. Two studies provided general lifestyle counselling focused on diet, exercise and weight management, without addiction-specific behavioural treatment[9,19]. No addiction-specific behavioural support was reported by 2 trials[14,18], while behavioural-support procedures were unclear or not reported in 3 further studies[10,11,20].

#### 2.5 Control

Across the 14 included trials, all comparator groups received placebo; no active pharmacological comparators were identified. Eleven studies used subcutaneously administered placebo controls, including matched placebo injections, saline preparations, sodium chloride solution or matching placebo pens[8-14,17,19-21]. Two semaglutide studies used a sham injection procedure involving a brief needle prick rather than an injected placebo substance[15,16]. Schacht et al[18] used a matched oral placebo. Comparator treatment durations ranged from 6 to 52 weeks. Five studies administered subcutaneous placebo daily to match liraglutide regimens[9,11,13,19,20], while Schacht et al[18] administered oral placebo daily. Eight studies used weekly injected placebo or sham procedures to match injectable semaglutide, exenatide or dulaglutide regimens[8,10,12,14-17,21].

#### 2.6 Outcomes

All 14 trials reported smoking and/or alcohol outcomes, with follow-up ranging from 6 to 60 weeks. Eight reported smoking outcomes[8,10,12,13,15-17,21], 11 reported alcohol outcomes[8-12,14,15,18-21] and 5 reported both[8,10,12,15,21]. Thirteen trials contributed to at least one meta-analysis; Fogelman et al[14] was narratively synthesised because sufficient numerical data were unavailable.

Four smoking outcomes were meta-analysed: smoking cessation, CPD, nicotine dependence and post-cessation weight. Smoking cessation was assessed in 3 studies using 7-day point-prevalence or prolonged abstinence[13,17,21]. Four studies reported cigarettes smoked per day[12,15,16,21], 3 reported Fagerström Test for Nicotine Dependence scores[8,10,12], and 2 provided post-cessation weight data[17,21]. Smoking-related outcomes were collected at follow-ups ranging from 6 to 52 weeks.

Nine alcohol outcomes were meta-analysed: daily alcohol consumption in grams[8,12,15,19], AUDIT scores[8-10,12,20], alcohol craving measured using the Penn Alcohol Craving Scale (PACS)[8,12,18], drinks per drinking day[12,15,18], drinks per calendar day[15,18], drinking days[15,18], drinks per week[9,11,21], heavy drinking days[8,12,15,18] and days without alcohol consumption[8,12,15]. WHO drinking-risk outcomes were synthesised narratively because sufficiently comparable data were unavailable for pooling[12,18].

Three studies biochemically verified smoking abstinence: Two used expired carbon monoxide[13,17], while 1 used carbon monoxide or urinary cotinine[21]. Other smoking outcomes were self-reported or questionnaire based. One trial[8] measured plasma phosphatidylethanol; the remaining alcohol outcomes relied principally on self-report.

Most final assessments occurred within 6-32 weeks of treatment initiation. Longer follow-up was reported by Armstrong et al[19] (60 weeks), Lüthi et al[21] ( 52 weeks) and Vedtofte et al[11] (52 weeks).

#### 2.7 List of Included Studies and Associated References

**eTable 2 List of included studies and associated references**

| **First author and year** | **Trial ID** | **Primary study reference, and other references** |
|---|---|---|
| Armstrong 2016 | NCT01237119<br><br>EudraCT 2009-016761-29 | **Primary:**<br>Armstrong MJ, Gaunt P, Aithal GP, Barton D, Hull D, Parker R, et al. Liraglutide safety and efficacy in patients with non-alcoholic steatohepatitis (LEAN): a multicentre, double-blind, randomised, placebo-controlled phase 2 study. The Lancet. 2016;387(10019):679–690. doi:10.1016/S0140-6736(15)00803-X.<br><br>**Secondary:**<br>University of Birmingham. A 48-week phase II, randomised, double-blinded, placebo-controlled, parallel-group, multi-centre trial on liraglutide's safety, efficacy and action on liver histology and metabolism in overweight patients. European Union Clinical Trials Register. EudraCT 2009-016761-29. Sponsor protocol HE2013. Registered 14 May 2010.<br><br>Armstrong M, Gaunt P, Aithal G, et al. OC-016 Liraglutide is effective in the histological clearance of non-alcoholic steatohepatitis in a multicentre, double-blinded, randomised, placebo-controlled phase II trial. Gut. 2015;64:A9. |
| Ashare 2018 | NCT03712098 | **Primary:**<br>Ashare RL. Daily liraglutide for nicotine dependence (DAL). ClinicalTrials.gov. NCT03712098; results posted September 2023. Accessed 1 May 2026.<br><br>**Secondary:**<br>Ashare RL, Amaro A. Glucagon-like peptide-1 receptor agonists as novel pharmacotherapies for nicotine dependence [clinical trial protocol]. Philadelphia (PA): Perelman School of Medicine, University of Pennsylvania; 2020. ClinicalTrials.gov. NCT03712098. Protocol version 8 (16 June 2020). |
| Fogelman 2025 | Potentially NCT04779697 | **Primary:**<br>Fogelman, N., Harvanek, Z., Horvath-Diano, C., Joseph, K., Doyle, E., Jastreboff, A., & Sinha, R. (2025). Semaglutide compared to placebo treatment decreases alcohol consumption and food craving over 12 weeks and a 1-month follow-up. Alcohol: Clinical and Experimental Research, 49. [Conference abstract]. |
| Hendershot 2026 | NCT05530577 | **Primary:**<br>Hendershot CS, Bremmer MP, Paladino MB, et al. Once-Weekly Semaglutide in Adults With Daily Cigarette Use: A Randomised Clinical Trial. JAMA Netw Open. 2026;9(5):e2614898. doi:10.1001/jamanetworkopen.2026.14898<br><br>**Secondary:** |

| | | |
|---|---|---|
| | | Hendershot CS, Jordan R, Tow A, Dermody S, Claus E, Klein K, Fletcher P. Effects of semaglutide on nicotine intake and smoking lapse [clinical trial protocol]. Protocol No. 21-1548. Version 1.7. 20 February 2023. |
| Hendershot 2025 | NCT05520775 | **Primary:**<br>Hendershot CS, Bremmer MP, Paladino MB, Kostantinis G, Gilmore TA, Sullivan NR, Tow AC, Dermody SS, Prince MA, Jordan R, McKee SA, Fletcher PJ, Claus ED, Klein KR. Once-weekly semaglutide in adults with alcohol use disorder: a randomised clinical trial. JAMA Psychiatry. 2025;82(4):395–405. doi:10.1001/jamapsychiatry.2024.4789.<br><br>**Secondary:**<br>Hendershot C. Semaglutide for alcohol use disorder. ClinicalTrials.gov. NCT05520775; results posted 2022. Accessed 1 May 2026.<br><br>Hendershot C. Human laboratory screening of semaglutide for alcohol use disorder [clinical trial protocol]. ClinicalTrials.gov. NCT05520775. Protocol No. 21-1689. Version 1.7. 9 February 2023. |
| Klausen 2022 | NCT03232112<br><br>EUCTR2016-003343-11-DK | **Primary:**<br>Klausen MK, Jensen ME, Møller M, Le Dous N, Jensen AMØ, Zeeman VA, et al. Exenatide once weekly for alcohol use disorder investigated in a randomised, placebo-controlled clinical trial. JCI Insight. 2022;7(19):e159863. doi:10.1172/jci.insight.159863.<br><br>**Secondary:**<br>Klausen MK, Jensen ME, Fink-Jensen A. The effects of exenatide once weekly for alcohol use disorder: results from a randomised, placebo-controlled clinical trial. Alcoholism: Clinical and Experimental Research. 2023;47(Suppl 1):65–66. doi:10.1111/acer.15069.<br><br>Does glucagon-like peptide (GLP-1) receptor agonist stimulation reduce alcohol intake in patients with alcohol dependence? ClinicalTrials.gov. 2017.<br><br>Jensen ME, Klausen MK, Bergmann ML, Knudsen GM, Vilsbøll T, Stove C, et al. Blood phosphatidylethanol measurements indicate GLP-1 receptor stimulation causes delayed decreases in alcohol consumption. Alcoholism: Clinical and Experimental Research. 2025;49(5):1161–1165. doi:10.1111/acer.70041.<br><br>European Union Clinical Trials Register. Does stimulation of the receptor for glucagon-like peptide-1 (GLP-1) reduce alcohol intake in patients with alcohol dependence. European Union Clinical Trials Register. EUCTR2016-003343-11-DK; 2017.<br><br>Antonsen KK, Klausen MK, Brunchmann AS, Le Dous N, Jensen ME, Miskowiak KW, et al. Does glucagon-like peptide-1 (GLP-1) receptor agonist stimulation reduce alcohol intake in patients with alcohol dependence: study protocol of a randomised, double-blinded, placebo-controlled clinical trial. BMJ Open. 2018;8(7):e019562. doi:10.1136/bmjopen-2017-019562. |
| Klausen 2026 | NCT05895643 | **Primary:**<br>Klausen MK, Justesen SK, Pedersen JN, Rasmussen L, Jensen A, Jensen ME, Knorr UB, Bergmann ML, Holst JJ, Hartmann B, Koob GF, Benveniste H, Volkow ND, Ekstrøm CT, Knudsen GM, Vilsbøll T, Fink-Jensen A. Once-weekly semaglutide versus placebo in patients |

| | | |
|---|---|---|
| | | with alcohol use disorder and comorbid obesity: a randomised, double-blind, placebo-controlled trial. The Lancet. 2026;407(10540):1687–1698. doi:10.1016/S0140-6736(26)00305-3.<br><br>**Secondary:**<br>Klausen MK, Kuzey T, Pedersen JN, Justesen SK, Rasmussen L, Knorr UB, Mason G, Ekstrom CT, Holst JJ, Koob G, Benveniste H, Volkow ND, Knudsen GM, Vilsbøll T, Fink-Jensen A. Does semaglutide reduce alcohol intake in Danish patients with alcohol use disorder and comorbid obesity? Trial protocol of a randomised, double-blinded, placebo-controlled clinical trial (the SEMALCO trial). BMJ Open. 2025;15(1):e086454. doi:10.1136/bmjopen-2024-086454.<br><br>ClinicalTrials.gov. Does semaglutide reduce alcohol intake in patients with alcohol use disorder and comorbid obesity? ClinicalTrials.gov. NCT05895643; 2023.<br><br>Clinical Trials Information System (CTIS). SEMALCO. Clinical Trials Information System. CTIS2023-503371-25-00; 2023.<br><br>Anonymous. Does the glucagon-like peptide 1 (GLP-1) receptor agonist semaglutide reduce alcohol intake in patients with alcohol use disorder and comorbid obesity? ClinicalTrials.gov. 2023. |
| Larsen 2017 | NCT01845259 | **Primary:**<br>Larsen JR, Vedtofte L, Jakobsen MSL, Jespersen HR, Jakobsen MI, Svensson CK, Koyuncu K, Schjerning O, Oturai PS, Kjaer A, Nielsen J, Holst JJ, Ekstrøm CT, Correll CU, Vilsbøll T, Fink-Jensen A. Effect of liraglutide treatment on prediabetes and overweight or obesity in clozapine- or olanzapine-treated patients with schizophrenia spectrum disorder: a randomised clinical trial. JAMA Psychiatry. 2017;74(7):719–728. doi:10.1001/jamapsychiatry.2017.1220.<br><br>Larsen JR, Vedtofte L, Holst JJ, Oturai P, Kjaer A, Correll CU, Vilsbøll T, Fink-Jensen A. Does a GLP-1 receptor agonist change glucose tolerance in patients treated with antipsychotic medications? Design of a randomised, double-blinded, placebo-controlled clinical trial. BMJ Open. 2014;4(3):e004227. doi:10.1136/bmjopen-2013-004227.<br><br>ClinicalTrials.gov. Does a GLP-1 receptor agonist change glucose tolerance in antipsychotic-treated patients? ClinicalTrials.gov. NCT01845259; 2013. |
| Lüthi 2024 | NCT03204396 | **Primary:**<br>Lüthi, H., Lengsfeld, S., Burkard, T., Meienberg, A., Jeanloz, N., Vukajlovic, T., ... & Winzeler, B. (2024). Effect of dulaglutide in promoting abstinence during smoking cessation: 12-month follow-up of a single-centre, randomised, double-blind, placebo-controlled, parallel group trial. EClinicalMedicine, 68. doi: 10.1016/j.eclinm.2024.102429<br><br>**Secondary:** |

| | | |
|---|---|---|
| | | Atila C, Burkard T, Meienberg A, Jeanloz N, Vukajlovic T, Bologna K, Steinmetz M, Sailer C, Coynel D, Vogt DR, Hemkens LG, Speich B, Kuhne J, Baur F, Lutz L, Zanchi D, Christ-Crain M, Winzeler B. Glucagon-like peptide-1 analogues: a new way to quit smoking? SKIP – a randomised controlled study. Journal of the Endocrine Society. 2022;6(Suppl 1):A261. doi:10.1210/jendso/bvac150.536.<br><br>Probst LS, Monnerat S, Lengsfeld S, Bathelt C, Meienberg A, Burkard T, Christ-Crain M, Winzeler BF. Effects of a GLP-1 agonist on consumption of alcohol in patients treated for smoking cessation. Journal of the Endocrine Society. 2023;7(Suppl 1):A643. doi:10.1210/jendso/bvad114.1200.<br><br>Probst L, Monnerat S, Vogt DR, Lengsfeld S, Burkard T, Meienberg A, Bathelt C, Christ-Crain M, Winzeler B. Effects of dulaglutide on alcohol consumption during smoking cessation. JCI Insight. 2023;8(22):e170419. doi:10.1172/jci.insight.170419.<br><br>ClinicalTrials.gov. Smoking cessation facilitated by glucagon-like peptide-1 (GLP-1) analogues. ClinicalTrials.gov. NCT03204396; 2017.<br><br>Anonymous. Smoking cessation facilitated by glucagon-like peptide-1 (GLP-1) analogues – a randomised, double-blind, placebo-controlled trial. ClinicalTrials.gov. 2017.<br><br>Baur F, Atila C, Lengsfeld S, Burkard T, Meienberg A, Bathelt C, Christ-Crain M, Winzeler B. Gender differences in weight gain during attempted and successful smoking cessation on dulaglutide treatment: a predefined secondary analysis of a randomised trial. BMJ Nutrition, Prevention and Health. 2023;6(2):301–309. doi:10.1136/bmjnph-2023-000781.<br><br>Lengsfeld S, Burkard T, Meienberg A, Jeanloz N, Coynel D, Vogt DR, Hemkens LG, Speich B, Zanchi D, Erlanger TE, Christ-Crain M, Winzeler B. Glucagon-like peptide-1 analogues: a new way to quit smoking? (SKIP) – a structured summary of a study protocol for a randomised controlled study. Trials. 2023;24(1):284. doi:10.1186/s13063-023-07164-9.<br><br>Baur F, Coynel D, Atila C, Lengsfeld S, Burkard T, Meienberg A, Bathelt C, Christ-Crain M, Winzeler BF. Gender differences in smoking cessation? Results of a randomised controlled trial of dulaglutide-assisted smoking cessation. Journal of the Endocrine Society. 2023;7(Suppl 1):A43–A44. doi:10.1210/jendso/bvad114.078.<br><br>Lengsfeld S, Burkard T, Meienberg A, Jeanloz N, Vukajlovic T, Bologna K, Steinmetz M, Bathelt C, Sailer CO, Vogt DR, Hemkens LG, Speich B, Urwyler SA, Kuhne J, Baur F, Lutz LN, Erlanger TE, Christ-Crain M, Winzeler B. Effect of dulaglutide in promoting abstinence during smoking cessation: a single-centre, randomised, double-blind, placebo-controlled, parallel group trial. eClinicalMedicine. 2023;57:101865. doi:10.1016/j.eclinm.2023.101865.<br><br>Sailer CO, Lengsfeld S, Coynel D, Baur F, Bologna K, Vukajlovic T, Jeanloz NA, Bathelt C, Zanchi D, Christ-Crain M, Winzeler BF. Functional and structural neuronal changes of dulaglutide compared to placebo during smoking cessation: a randomised placebo-controlled trial. Journal of the Endocrine Society. 2024;8(Suppl 1):A634–A635. doi:10.1210/jendso/bvae163.1217. |

| | | |
|---|---|---|
| | | Beck J, Hasenbohler F, Werlen L, Lengsfeld S, Meienberg A, Bathelt C, Vogt D, Christ-Crain M, Burkard T, Winzeler B. Blood pressure changes during smoking cessation in a randomised, double-blind, placebo-controlled trial of dulaglutide treatment. European Journal of Preventive Cardiology. 2025. doi:10.1093/eurjpc/zwaf055. |
| Miras 2019 | ISRCTN13643081 | **Primary:**<br>Miras AD, Pérez-Pevida B, Aldhwayan M, Kamocka A, McGlone ER, Al-Najim W, Chahal H, Batterham RL, McGowan B, Khan O, Greener V, Ahmed AR, Petrie A, Scholtz S, Bloom SR, Tan TM. Adjunctive liraglutide treatment in patients with persistent or recurrent type 2 diabetes after metabolic surgery (GRAVITAS): a randomised, double-blind, placebo-controlled trial. The Lancet Diabetes & Endocrinology. 2019;7(7):549–559. doi:10.1016/S2213-8587(19)30157-3.<br><br>**Secondary:**<br>ISRCTN Registry. The effects of liraglutide in controlling blood sugar and weight in poor-responders to bariatric surgery. ISRCTN Registry. ISRCTN13643081; 2016. |
| Sass, 2026 | NCT04892199 | **Primary:**<br>Sass MR, Klausen MK, Schwarz CR, Rasmussen L, Giver MEB, Hviid M, Schilling C, Zamorski A, Jensen A, Gefke M, Storgaard H, Oturai PS, Kjaer A, Hartmann B, Holst JJ, Ekstrøm CT, Vinberg M, Correll CU, Vilsbøll T, Fink-Jensen A. Semaglutide and early-stage metabolic abnormalities in individuals with schizophrenia spectrum disorders: a randomised clinical trial. JAMA Psychiatry. 2026;83(2):128–138. doi:10.1001/jamapsychiatry.2025.3639.<br><br>**Secondary:**<br>ClinicalTrials.gov. Does GLP-1RA prevent deterioration of metabolic state in prediabetic and diabetic patients treated with antipsychotic medication? ClinicalTrials.gov. NCT04892199; 2021.<br><br>Sass MR, Danielsen AA, Kohler-Forsberg O, Storgaard H, Knop FK, Nielsen MO, Sjodin AM, Mors O, Correll CU, Ekstrom C, Vinberg M, Nielsen J, Vilsbøll T, Fink-Jensen A. Effect of the GLP-1 receptor agonist semaglutide on metabolic disturbances in clozapine-treated or olanzapine-treated patients with a schizophrenia spectrum disorder: study protocol of a placebo-controlled, randomised clinical trial (SemaPsychiatry). BMJ Open. 2023;13(1):e068652. doi:10.1136/bmjopen-2022-068652.<br><br>Anonymous. Does the glucagon-like peptide-1 receptor agonist semaglutide prevent deterioration of metabolic state in prediabetic or diabetic patients with schizophrenia treated with the antipsychotic compounds clozapine or olanzapine? ClinicalTrials.gov. 2021.<br><br>European Union Clinical Trials Register. Does the glucagon-like peptide-1 receptor agonist semaglutide prevent deterioration of metabolic state in prediabetic or diabetic patients with schizophrenia treated with the antipsychotic compounds clozapine or olanzapine? European Union Clinical Trials Register. EUCTR2020-000102-28-DK; 2020. |

| | | |
|---|---|---|
| Schacht 2026 | clinicaltrials.gov, NCT05892432 | **Primary:**<br>Schacht JP, Sakai JT, Raymond K, Shelton R. Oral Semaglutide for Alcohol Use Disorder: A Randomised Clinical Trial. Am J Psychiatry. 2026 Sep 1;183(9):636-645. doi: 10.1176/appi.ajp.20260003. Epub 2026 Jul 29.<br><br>**Secondary:**<br>ClinicalTrials.gov (2023). *Clinical trial of Rybelsus (semaglutide) among adults with alcohol use disorder (AUD).* NCT05892432.<br><br>Schacht, J. P., Sakai, J. T., Raymond, K., & Shelton, R. (2026). Oral semaglutide for alcohol use disorder: A randomised clinical trial. *American Journal of Psychiatry*. https://doi.org/10.1176/appi.ajp.20260003 |
| Vedtofte 2020 | EUCTR2012-001371-37-DK | **Primary:**<br>Vedtofte L, Bahne E, Foghsgaard S, Bagger JI, Andreasen C, Strandberg C, Gørtz PM, Holst JJ, Grønbæk H, Svare JA, Clausen TD, Mathiesen ER, Damm P, Gluud LL, Knop FK, Vilsbøll T. One year's treatment with the glucagon-like peptide 1 receptor agonist liraglutide decreases hepatic fat content in women with nonalcoholic fatty liver disease and prior gestational diabetes mellitus in a randomised, placebo-controlled trial. Journal of Clinical Medicine. 2020;9(10):3213. doi:10.3390/jcm9103213.<br><br>**Secondary:**<br>European Union Clinical Trials Register. The impact of liraglutide on glucose tolerance and the risk of type 2 diabetes in women with previous gestational diabetes mellitus. European Union Clinical Trials Register. EUCTR2012-001371-37-DK; 2012.<br><br>Foghsgaard S, Vedtofte L, Mathiesen ER, Svare JA, Gluud LL, Holst JJ, Damm P, Knop FK, Vilsbøll T. The effect of a glucagon-like peptide-1 receptor agonist on glucose tolerance in women with previous gestational diabetes mellitus: protocol for an investigator-initiated, randomised, placebo-controlled, double-blinded, parallel intervention trial. BMJ Open. 2013;3(10):e003834. doi:10.1136/bmjopen-2013-003834. |
| Yammine 2021 | NCT02975297 | **Primary:**<br>Yammine L, Green CE, Kosten TR, de Dios C, Suchting R, Lane SD, Verrico CD, Schmitz JM. Exenatide adjunct to nicotine patch facilitates smoking cessation and may reduce post-cessation weight gain: a pilot randomised controlled trial. Nicotine and Tobacco Research. 2021;23(10):1682–1690. doi:10.1093/ntr/ntab066.<br><br>**Secondary:**<br>Yammine L, de Dios C, Suchting R, Green CE, Nielsen DA, Walss-Bass C, Schmitz JM. Exploring predictors of treatment response to GLP-1 receptor agonists for smoking cessation. Nicotine and Tobacco Research. 2025;27(7):1294–1300. doi:10.1093/ntr/ntaf005.<br><br>Anonymous. Exenatide once weekly for smoking cessation: a randomised clinical trial. ClinicalTrials.gov. 2016. |

| | | |
|---|---|---|
| | | Yammine L, Kosten TR, Cinciripini PM, Green CE, Meininger JC, Minnix JA, Newton TF. Exenatide once weekly for smoking cessation: study protocol for a randomised clinical trial. Medicine. 2018;97(2):e9567. doi:10.1097/MD.0000000000009567. |
| **Abbreviations:** European Union Drug Regulating Authorities Clinical Trials (EudraCT); EU Clinical Trials Register (EUCTR); International Standard Randomised Controlled Trial (ISRCT) registry; NCT (National Clinical Trials) registry (a.k.a. ClinicalTrials.gov) | | |

#### 2.8 Further Characteristics of Included Studies

**eTable 3 Trial design, funding, conflicts of interest and eligibility characteristics of included studies**

| Study | Trial registry (ID) | Publication status | Trial design | Key eligibility criteria | Funding source(s) | Author declarations / COIs |
|---|---|---|---|---|---|---|
| Armstrong (2016) | NCT01237119 | Published | Double-blind placebo-controlled RCT | Adults with NASH and BMI >=25 kg/m2; significant non-NASH liver disease excluded. | Wellcome Trust; NIHR; Novo Nordisk | Multiple authors reported honoraria, advisory roles, or trial support from Novo Nordisk and other pharmaceutical companies. |
| Ashare (2018) | NCT03712098 | Unpublished (registry results) | Double-blind placebo-controlled RCT | Smokers with overweight/obesity seeking cessation treatment; major psychiatric illness excluded. | Novo Nordisk Inc. | Not reported. |
| Fogelman (2025) | NCT04779697 (potentially) | Conference abstract | Randomised placebo-controlled RCT; secondary alcohol analysis | Adults with obesity who regularly consumed alcohol; recruited to an obesity/food-reward trial rather than specifically for alcohol treatment. | NIDDK/NIH; Yale University (parent trial; provisional pending confirmation of trial linkage) | Not reported in abstract. |
| Hendershot (2026) | NCT05530577 | Published | Phase 2a, triple-blind, placebo-controlled parallel RCT | Non-treatment-seeking adults aged 21–65 who smoked ≥5 cigarettes/day (CO ≥8 ppm); key exclusions included diabetes, BMI <23 kg/m², recent cessation treatment/NRT, regular e-cigarette use, and other substance use disorders. | National Institute on Drug Abuse (NIDA); NCATS/NIH support | Hendershot reported personal fees from Eli Lilly and Apollo Therapeutics; McKee reported NIH grants and Elsevier fees; Klein reported fees/research support from Novo Nordisk and multiple other pharmaceutical companies. |
| Hendershot (2025) | NCT05520775 | Published | Randomised placebo-controlled trial | Non-treatment-seeking adults with AUD and hazardous drinking; severe withdrawal risk excluded. | NIAAA; NCATS; University of North Carolina | One author reported consultancy fees and institution-level contracts with pharmaceutical companies. |
| Klausen (2022) | NCT03232112 | Published | Double-blind placebo-controlled RCT | Treatment-seeking adults with AUD. | Novavi Foundation; Capital Region of Denmark; Novo Nordisk medication support | Several authors reported relationships with Novo Nordisk and other pharmaceutical companies. |

| | | | | | | |
|---|---|---|---|---|---|---|
| Klausen (2026) | NCT05895643 | Published | Double-blind placebo-controlled RCT | Treatment-seeking adults with AUD and obesity; severe psychiatric illness excluded. | Capital Region of Denmark; Novo Nordisk; Lundbeck Foundation; Novo Nordisk Foundation | Multiple authors reported consultancy, advisory board memberships, grants, and research support from pharmaceutical companies. |
| Larsen (2017) | NCT01845259 | Published | Double-blind placebo-controlled RCT | Adults with schizophrenia-spectrum disorders receiving clozapine /olanzapine and prediabetes/obesity. | Novo Nordisk; Lundbeck Foundation; Capital Region Psychiatry Research Group | Multiple authors reported speaker fees, consultancy, grants, and advisory roles with pharmaceutical companies, including Novo Nordisk. |
| Lüthi (2024) | NCT03204396 | Published | Double-blind placebo-controlled RCT | Treatment-seeking smokers with FTND >=5. | Swiss National Science Foundation; Bangerter-Rhyner Foundation; Swiss Heart Foundation | One author reported unrelated Moderna funding; otherwise no major study-related COIs. |
| Miras (2019) | ISRCTN13643081 | Published | Double-blind placebo-controlled RCT | Adults with obesity and recurrent/persistent T2D following bariatric surgery. | NIHR; MRC; BBSRC; Novo Nordisk; JP Moulton Foundation; EU FP7 | Multiple authors reported grants, honoraria, consultancy, and trial support from Novo Nordisk and other industry partners. |
| Sass (2026) | NCT04892199 | Published | Double-blind placebo-controlled RCT | Adults with schizophrenia-spectrum disorders and early glycemic dysregulation receiving clozapine/olanzapine. | Mental Health Services Capital Region Denmark; Novo Nordisk; Lundbeck Foundation; Novo Nordisk Foundation | Extensive industry relationships reported. |
| Schacht 2026 | NCT05892432 | Published | Phase 2, randomised, placebo-controlled parallel RCT | Treatment-seeking adults ≥21 years with moderate/severe AUD, BMI ≥25 kg/m² and high alcohol consumption; key exclusions included current psychiatric disorder, suicidality, moderate/severe non-alcohol substance use disorder, recent AUD/weight-loss medication and previous GLP-1RA use. | NIAAA; NCATS/Colorado Clinical and Translational Sciences Institute; Hewit Family Foundation; anonymous family donor | Schacht reported advisory/consultancy roles with Apollo Therapeutics and SOURCE Bio, Altimmune trial funding/study role, study medication from Bausch Health, and membership of an initiative supported by Eli Lilly and Imbrium Therapeutics; other authors declared no commercial financial relationships. |

| | | | | | | |
|---|---|---|---|---|---|---|
| Vedtofte (2020) | EUCTR2012-001371-37 | Published | Double-blind placebo-controlled RCT | Women with previous gestational diabetes and overweight/obesity; current diabetes excluded. | Novo Nordisk; Novo Nordisk Foundation; Danish Diabetes Academy; charitable foundations | Several authors reported advisory board membership, honoraria, grants, or industry collaborations. |
| Yammine (2021) | NCT02975297 | Published | Double-blind placebo-controlled RCT | Smokers with prediabetes and/or overweight; current cessation treatment excluded. | University of Texas Health Science Center at Houston | No conflicts of interest declared. |

**Abbreviations:** AUD, alcohol use disorder; BMI, body mass index; BBSRC, Biotechnology and Biological Sciences Research Council; COI, conflict of interest; EU FP7, European Union Seventh Framework Programme; FTND, Fagerström Test for Nicotine Dependence; NASH, non-alcoholic steatohepatitis; NCATS, National Center for Advancing Translational Sciences; NIAAA, National Institute on Alcohol Abuse and Alcoholism; NIDA, National Institute on Drug Abuse; NIHR, National Institute for Health and Care Research; MRC, Medical Research Council; RCT, randomised controlled trial; T2D, type 2 diabetes.

##### 2.9 Ongoing Studies

We identified 30 ongoing studies or completed studies awaiting results. Fourteen will assess alcohol-related outcomes, 8 will assess tobacco-related outcomes, and 4 will assess both alcohol and tobacco outcomes. Four indicate they are trials for addiction in the title, but do not report a long list of outcomes in the registry. These studies will evaluate outcomes including alcohol consumption, heavy drinking, abstinence, craving and alcohol biomarkers, as well as smoking cessation or reduction, nicotine dependence, craving, withdrawal, carbon monoxide levels and post-cessation weight. Full details are provided in the Characteristics of ongoing studies (**eTable 4**).

#### eTable 4 Registered, ongoing, completed and unpublished trials investigating GLP-1 receptor agonists for smoking and alcohol-related outcomes

| Trial registry | Recruitment status | Funding | Country | Population | Intervention | Tobacco- and/or alcohol-related outcomes | Start date | End date |
|---|---|---|---|---|---|---|---|---|
| ACTRN12626000530325 | Not yet recruiting | Australian Government Department of Health, Disability and Ageing, Medical Research Future Fund (MRFF); sponsor: The University of Queensland. | Australia | Adults with schizophrenia or schizoaffective disorder and overweight or obesity. | Tirzepatide, titrated to 10 mg weekly for 24 weeks, versus saline placebo; routine care continued in both groups. | Nicotine dependence (FTND). | Jun 2026 | Jun 2029 |
| jRCTs061250103 | Recruiting | No research funding reported; sponsor: Okayama University Hospital. | Japan | Adults aged 20–65 years with alcohol dependence and high- or very-high-risk drinking. | Tirzepatide, titrated to 5–7.5 mg weekly for 12 weeks, versus oral nalmefene 10–20 mg before drinking. | Heavy drinking days; total alcohol consumption; WHO drinking-risk level; alcohol craving. | Mar 2026 | Mar 2030 |
| NCT05610800 | Not recruiting | The University of Texas Health Science Center, Houston. | USA | Adults who smoke and are seeking smoking cessation treatment. | Exenatide 2 mg versus placebo; both groups received nicotine patches and cessation counselling. | Biochemically verified continuous smoking abstinence; post-cessation weight. | Not reported | Jan 2026 |
| NCT05891587 | Completed; results unpublished | Oklahoma State University Center for Health Sciences. | USA | Adults with alcohol use disorder and heavy drinking. | Semaglutide, titrated from 0.25 to 1.0 mg weekly for 12 weeks, versus saline placebo. | Drinks per drinking day and week; heavy drinking days; PEth. | Jul 2023 | Oct 2025 |
| NCT06004830 | Completed | Yale University (funding not reported). | USA | Adults with HIV, alcohol use disorder, PEth greater than 20 ng/mL, and polypharmacy. | Spironolactone with pharmacist and addiction-psychiatrist support; no comparator was reported. | Self-reported alcohol use; PEth. | Nov 2023 | Jan 2026 |
| NCT06006143 | Active; not recruiting | Yale University (funding not reported). | USA | Adults with HIV and alcohol use disorder who currently smoke cigarettes. | Varenicline with pharmacist and addiction-psychiatrist support; no comparator was reported. | Self-reported alcohol use; PEth. | Jun 2025 | Aug 2026 |
| NCT06015893 | Recruiting | University of North Carolina, Chapel Hill. | USA | Treatment-seeking adults with alcohol use disorder and heavy drinking. | Dose-escalated semaglutide versus matched placebo. | Alcohol consumption; heavy drinking days; alcohol craving. | Aug 2023 | Dec 2026 |
| NCT06173778 | Recruiting | The University of Texas Health Science Center, Houston. | USA | Adults with overweight or obesity who smoke and are attempting cessation. | Semaglutide 2.4 mg weekly versus placebo; both groups received nicotine patches and brief cessation counselling. | Smoking abstinence; post-cessation weight. | Apr 2024 | Aug 2026 |
| NCT06409130 | Completed; results unpublished | Novo Nordisk A/S. | Multinational | Adults with alcohol-related liver disease. | Semaglutide, cagrilintide, NNC0194-0499, or cagrilintide plus semaglutide versus matched placebo. | Alcohol consumption (g/day); PEth. | May 2024 | Jan 2026 |
| NCT06546384 | Not yet recruiting | Insel Gruppe AG, University Hospital Bern. | Switzerland | Adults with overweight or obesity, fatty liver disease, and hazardous alcohol use or alcohol use disorder. | Semaglutide, titrated to 2.4 mg weekly, plus weight-management support versus weight-management support alone. | Alcohol abstinence; alcohol-free, drinking and heavy drinking days; drinks/week; PEth and EtG. | Jun 2025 | Apr 2027 |
| NCT06727331 | Recruiting | Brigham and Women's Hospital. | USA | Adults with alcohol use disorder. | Tirzepatide versus saline placebo; the dose was not reported. | Cue-induced alcohol craving; PACS. | Sep 2025 | May 2026 |
| NCT06924697 | Enrolling | Academic Leaders Training Program of Pudong Health Bureau of Shanghai; Special Care Program for Obesity in Pudong Health Bureau of Shanghai. | China | Men aged 18–75 years with type 2 diabetes who currently smoke and have FTND scores of 4 or higher. | Routine GLP-1 receptor agonist therapy, mainly semaglutide or liraglutide, versus a DPP-4 inhibitor. | Nicotine dependence (FTND); exhaled carbon monoxide; smoking cessation. | Jun 2025 | Jul 2026 |
| NCT06939088 | Recruiting | Anders Fink-Jensen, MD, DMSci (funding not reported). | Denmark | Adults aged 18–70 years with alcohol dependence, schizophrenia-spectrum disorder, AUDIT scores above 15, and BMI of 23 kg/m² or higher. | Once-weekly tirzepatide versus saline placebo. | Heavy drinking and alcohol-free days; total consumption; WHO drinking-risk level; PACS; AUDIT; nicotine dependence. | May 2025 | Dec 2028 |
| NCT06986993 | Recruiting | University of Oklahoma. | USA | Adults with obesity who smoke daily and are not currently trying to quit. | Once-weekly semaglutide versus placebo; the dose was not reported. | Smoking behaviour/reduction; nicotine craving and withdrawal; motivation to quit. | Jul 2024 | May 2028 |
| NCT06987513 | Completed; results unpublished | Altimmune, Inc. (funding not reported). | USA | Adults with moderate or severe alcohol use disorder and overweight or obesity. | Pemvidutide 2.4 mg weekly versus placebo. | Heavy drinking days; WHO drinking-risk level; PEth; drinks per drinking day; abstinent days. | May 2025 | Jul 2026 |

| | | | | | | | | |
|---|---|---|---|---|---|---|---|---|
| NCT06994338 | Recruiting | Keck School of Medicine, University of Southern California Funder type: U.S. National Institutes of Health (grant listed in study IDs). | USA | Adults with alcohol use disorder, including people with overweight or obesity. | Tirzepatide versus placebo; the dosing schedule was not reported. | Heavy drinking days; alcohol consumption and related behavioural outcomes. | Oct 2025 | Aug 2026 |
| NCT07040592 | Recruiting | Yale University (funding not reported). | USA | Adults with HIV, alcohol use disorder, PEth greater than 20 ng/mL, and polypharmacy. | Semaglutide with pharmacist and addiction-psychiatrist support; no comparator was reported. | Self-reported alcohol use; PEth. | Jan 2026 | Jun 2027 |
| NCT07046819 | Not yet recruiting | National Institute on Alcohol Abuse and Alcoholism (NIAAA). | USA | Adults with alcohol use disorder, metabolic alcohol-associated liver disease, and BMI of 25 to less than 40 kg/m². | Tirzepatide, titrated to 7.5 mg weekly for 12 weeks, versus saline placebo. | Drinking behaviour and craving; WHO drinking-risk level. | Jun 2026 | Jul 2027 |
| NCT07059377 | Not yet recruiting | Ottawa Heart Institute Research Corporation. | Canada | Adults with type 2 diabetes who smoke at least 5 cigarettes daily and are willing to reduce or quit. | Semaglutide, titrated to 2 mg weekly for up to 26 weeks, plus nicotine replacement therapy versus nicotine replacement therapy alone. | Smoking cessation at 6 months. | Feb 2026 | Jul 2027 |
| NCT07136714 | Recruiting | Nordsjaellands Hospital (funding not reported). | Denmark | Adults aged 18–65 years with depression and BMI of 27 kg/m² or higher. | Once-weekly semaglutide versus saline placebo. | Nicotine dependence (measure and timing not specified in extraction). | Sep 2025 | Sep 2027 |
| NCT07218354 | Recruiting | VA Office of Research and Development (funding not reported). | USA | Veterans with moderate or severe alcohol use disorder and high- or very-high-risk drinking who want to reduce consumption. | Semaglutide, titrated to 2.4 mg weekly or the maximum tolerated dose, versus placebo. | WHO drinking-risk level; no heavy drinking days. | Jul 2026 | Mar 2030 |
| NCT07221214 | Recruiting | Vanderbilt University Medical Center (funding not reported). | USA | Adults with HIV receiving stable antiretroviral therapy and with BMI of 23 kg/m² or higher. | Oral semaglutide versus placebo. | Drinks/week; cigarettes/day. | Apr 2026 | Jan 2030 |
| NCT07223983 | Recruiting | Yale University (funding not reported). | USA | Adults with alcohol use disorder and obesity or overweight after bariatric surgery. | Semaglutide, titrated to 1 mg weekly for 3 months; the comparator was not reported. | Drinks per calendar and drinking day; heavy drinking, drinking and abstinent days. | Mar 2026 | Oct 2026 |
| NCT07227948 | Recruiting | The University of Texas Health Science Center, Houston (funding not reported). | USA | Adults with BMI of 25 kg/m² or higher enrolled in a cannabis-use disorder trial. | Semaglutide, titrated to 1 mg weekly for 14 weeks, versus saline placebo; both groups received cognitive behavioural therapy. | Tobacco/nicotine and alcohol use (secondary measures not specified in extraction). | Jan 2026 | Feb 2029 |
| NCT07249554 | Not yet recruiting | Johns Hopkins University (funding not reported). | USA | Adults aged 21–65 years receiving residential treatment for alcohol use disorder. | An oral GLP-1 receptor agonist, oral naltrexone, or placebo. | Alcohol-use outcomes (preliminary efficacy measures not specified in extraction). | Sep 2026 | Sep 2028 |
| NCT07292519 | Not yet recruiting | South West Sydney Local Health District (funding not reported). | Australia | Adults aged 21–75 years with moderate or severe alcohol use disorder, high alcohol consumption, and BMI of 27 kg/m² or higher. | Tirzepatide, titrated to 5 mg weekly for 8 weeks, versus placebo; both groups received a digital cognitive behavioural intervention. | Heavy drinking days; drinks per drinking day; abstinent days; craving; WHO drinking-risk level. | Jan 2026 | Jan 2028 |
| NCT07509112 | Not yet recruiting | Kirby Institute (funding not reported). | Australia | Adults with moderate or severe methamphetamine use disorder. | Semaglutide, titrated to 1 mg weekly for 12 weeks; the comparator was not reported. | Tobacco/nicotine and alcohol use (exploratory measures not specified in extraction). | Sep 2026 | Mar 2027 |
| NCT07523633 | Recruiting | Anders Fink-Jensen, MD, DMSci (funding not reported). | Denmark | Adults aged 18–70 years seeking to reduce or stop cannabis use and with BMI of 23 kg/m² or higher. | Semaglutide, titrated to 2.4 mg weekly, versus saline placebo. | Severity of alcohol use and nicotine dependence. | Apr 2026 | Apr 2028 |
| NCT07559500 | Recruiting | National Institute on Alcohol Abuse and Alcoholism (NIAAA) (funding not reported). | USA | Adults with prior stimulant exposure, including a cohort with mild or moderate alcohol use disorder and heavy drinking. | Tirzepatide versus saline placebo, with PET imaging to assess dopamine signalling. | Alcohol craving and withdrawal symptoms. | May 2026 | Dec 2031 |
| NCT07602699 | Recruiting | University of Southern California (funding not reported). | USA | Adults with overweight or obesity who smoke at least 10 cigarettes daily and intend to quit. | Tirzepatide, titrated to 10 mg weekly for 16 weeks, versus saline placebo. | 7-day point-prevalence smoking abstinence; cigarettes/day; craving. | Jun 2026 | Sep 2028 |
| Abbreviations: AUD, alcohol use disorder; AUDIT, Alcohol Use Disorders Identification Test; EtG, ethyl glucuronide; FTND, Fagerström Test for Nicotine Dependence; PACS, Penn Alcohol Craving Scale; PEth, phosphatidylethanol; WHO, World Health Organization. Registry information reflects the supplied extraction completed in September 2026 and may change. | | | | | | | | |

#### 2.10 Studies Excluded at Full-Text Review

**eTable 5 Sample of studies excluded at full-text review**

| Reference | Primary exclusion reason |
|---|---|
| Astrup A., Carraro R., Finer N., Harper A., Kunesova M., Lean M.E.J., Niskanen L., Rasmussen M.F., Rissanen A., Rossner S., Savolainen M.J., & Van Gaal L. (2012). Safety, tolerability and sustained weight loss over 2 years with the once-daily human GLP-1 analog, liraglutide. International Journal of Obesity, 36(6), 843-854. https://doi.org/10.1038/ijo.2011.158 | Wrong outcomes |
| Bonaca, M. P., Catarig, A. M., Hansen, Y., Houlind, K., Ramesh, C. K., Ludvik, B., Nordanstig, J., Rasouli, N., Sourij, H., & Verma, S. (2025). Design and baseline characteristics of the STRIDE trial: evaluating semaglutide in people with symptomatic peripheral artery disease and type 2 diabetes. European heart journal. Cardiovascular pharmacotherapy, 10(8), 728-737. https://doi.org/10.1093/ehjcvp/pvae071 | Wrong outcomes |
| Lofton H.F., Maranga G., Hold R., Fielding G., Youn H., Gujral A., Heffron S., & Fielding C. (2025). A randomised, double-blind, placebo-controlled trial of weight loss using liraglutide 3.0 mg for weight recurrence after Roux-en-Y gastric bypass. Surgery for Obesity and Related Diseases, 21(2), 135-145. https://doi.org/10.1016/j.soard.2024.08.037 | Wrong outcomes |
| Maddaloni E., Nguyen M., Shah S.H., & Holman R.R. (2025). Osteoprotegerin, Osteopontin, and Osteocalcin Are Associated With Cardiovascular Events in Type 2 Diabetes: Insights From EXSCEL. Diabetes Care, 48(2), 235-242. https://doi.org/10.2337/dc24-1455 | Wrong outcomes |
| Mahalingam S., Bellamkonda R., Yoon J., Perumal S.K., Casey C., Kharbanda K., & Rasineni K. (2023). Glucagon-like peptide 1 receptor agonist, Exendin-4, reduces alcohol-associated fatty liver disease. Alcoholism: Clinical and Experimental Research, 47(Supplement 1), 502. https://doi.org/10.1111/acer.5071 | Wrong outcomes |
| Mimura H., Oura T., Chin R., Takeuchi M., Fujihara K., & Sone H. (2025). Association of bodyweight loss with changes in lipids, blood pressure, and fasting serum glucose following tirzepatide treatment in Japanese participants with type 2 diabetes: A post hoc analysis of the SURPASS J-mono trial. Journal of Diabetes Investigation, 16(5), 807-816. https://doi.org/10.1111/jdi.14395 | Wrong outcomes |
| Mori K., Hamamoto Y., Honjo S., Kawasaki Y., Tatsuoka H., Fujimoto K., Matsuoka A., Ikeda H., Wada Y., & Koshiyama H. (2012). Comparison of liraglutide and exenatide evaluated by Continuous Glucose Monitoring (CGM) in patients with diabetes associated with liver cirrhosis. Diabetes, 61(SUPPL. 1), A292-A293. https://doi.org/10.2337/db12-836-1328 | Wrong outcomes |
| Satman I. (2015). LEADER-6: Variation of renal function status and risk markers across geographic origin, race and ethnicity in a high CV risk type 2 diabetic population: Baseline results of the trial. Diabetologia, 58(1 SUPPL. 1), S85. https://doi.org/10.1007/s00125-015-3687-4 | Wrong outcomes |
| Verma S., Leiter L.A., David Mazer C., Bain S.C., Buse J., Marso S., Nauck M., Zinman B., Bosch-Traberg H., Rasmussen S., Michelsen M.M., & Bhatt D.L. (2018). Liraglutide reduces cardiovascular events and mortality in type 2 diabetes mellitus independently of baseline low-density lipoprotein cholesterol levels and statin use results from the LEADER trial. Circulation, 138(15), 1605-1607. https://doi.org/10.1161/CIRCULATIONAHA.118.036862 | Wrong outcomes |
| Wijkman M.O., Claggett B., Diaz R., Gerstein H.C., Kober L., Lewis E., Maggioni A.P., Wolsk E., Aguilar D., Bentley-Lewis R., McMurray J.J., Probstfield J., Riddle M., Tardif J.-C., Solomon S.D., & Pfeffer M.A. (2020). Blood pressure and mortality in patients with type 2 diabetes and a recent coronary event in the ELIXA trial. Cardiovascular Diabetology, 19(1), 175. https://doi.org/10.1186/s12933-020-01150-0 | Wrong outcomes |

| | |
|---|---|
| Farokhnia, M., Tazare, J., Pince, C., Bruns, N., Gray, J., Lo Re, V., Fiellin, D., Kranzler, H., Koob, G., Justice, A., Vendruscolo, L., Rentsch, C., & Leggio, L. (2024). GLUCAGON-LIKE PEPTIDE-1 RECEPTOR AGONISTS BUT NOT DIPEPTIDYL PEPTIDASE-4 INHIBITORS REDUCE ALCOHOL INTAKE: CONVERGING EVIDENCE FROM RODENT BEHAVIORAL PHARMACOLOGY AND HUMAN PHARMACOEPIDEMIOLOGIC STUDIES. NEUROPSYCHOPHARMACOLOGY, 49, 289-290. | Wrong study design |
| Hardonova M., Siarnik P., Sivakova M., Sucha B., Penesova A., Radikova Z., Havranova A., Imrich R., Vlcek M., Zitnanova I., Krastev G., Kiacikova M., Kollar B., & Turcani P. (2023). Endothelial Function in Patients with Multiple Sclerosis: The Role of GLP-1 Agonists, Lipoprotein Subfractions, and Redox Balance. International Journal of Molecular Sciences, 24(13), 11162. https://doi.org/10.3390/ijms241311162 | Wrong study design |
| Knop F.K., Harring S., Holst I., Kvist K., Lingvay I., & Vilsboll T. (2020). Gastrointestinal adverse events with once-weekly semaglutide: Risk predictors and effect on semaglutide response. Diabetologia, 63(SUPPL 1), S290-S291. https://doi.org/10.1007/s00125-020-05221-5 | Wrong study design |
| Quddos F., Fowler M., de Lima Bovo A.C., Elbash Z., Tegge A.N., Gatchalian K.M., Kablinger A.S., Di Feliceantonio A.G., & Bickel W.K. (2025). Physiological and perceptual effects of GLP-1 receptor agonists during alcohol consumption in people with obesity: a pilot study. medRxiv. https://doi.org/10.1101/2025.04.25.25326413 | Wrong study design |
| Yella, S., Sree, K., & Mahato, S. (2025). The Role of Glucagon-Like Peptide-1 Receptor Agonists in the Treatment of Alcohol Use Disorder. JOURNAL OF CLINICAL PSYCHOPHARMACOLOGY, 45(4), 372-375. https://doi.org/10.1097/jcp.000000000000201010.1097/JCP.0000000000002010 | Wrong study design |
| Cooley B.J., Occelli Hanbury-Brown C.V., Choi E.A., Heller W.A., Lim A.W., Lawrence A.J., Haber P.S., McNally G.P., & Millan E.Z. (2025). FGF21 analogue PF-05231023 on alcohol consumption and neuronal activity in the nucleus accumbens. Neuropsychopharmacology, e8740. https://doi.org/10.1038/s41386-025-02133-z | Wrong intervention |
| Davis J.F., Schurdak J.D., Magrisso I.J., Mul J.D., Grayson B.E., Pfluger P.T., Tschop M.H., Seeley R.J., & Benoit S.C. (2012). Gastric bypass surgery attenuates ethanol consumption in ethanol-preferring rats. Biological Psychiatry, 72(5), 354-360. https://doi.org/10.1016/j.biopsych.2012.01.035 | Wrong intervention |
| Haass-Koffler C.L., Giovenco D.E., Lee M.R., Zywiak W.H., De La Monte S.M., Kenna G.A., Swift R.M., & Leggio L. (2016). Serum insulin levels are reduced by intravenous ghrelin administration but do not correlate with alcohol craving in alcohol-dependent individuals. International Journal of Neuropsychopharmacology, 19(10), 1-7. https://doi.org/10.1093/ijnp/pyw048 | Wrong intervention |
| Lee M.R., Farokhnia M., Cobbina E., Saravanakumar A., Li X., Battista J.T., Farinelli L.A., Akhlaghi F., & Leggio L. (2020). Endocrine effects of the novel ghrelin receptor inverse agonist PF-5190457: Results from a placebo-controlled human laboratory alcohol co-administration study in heavy drinkers. Neuropharmacology, 170, 107788. https://doi.org/10.1016/j.neuropharm.2019.107788 | Wrong intervention |
| Sharma A.N., Pise A., Sharma J.N., & Shukla P. (2015). Dipeptidyl-peptidase IV (DPP-IV) inhibitor delays tolerance to anxiolytic effect of ethanol and withdrawal-induced anxiety in rats. Metabolic Brain Disease, 30(3), 659-667. https://doi.org/10.1007/s11011-014-9603-7 | Wrong intervention |
| Armstrong, M., Hull, D., Guo, K., Barton, D., Yu, J., Tomlinson, J., & Newsome, P. (2014). Effect of liraglutide on adipose insulin resistance and hepatic de-novo lipogenesis in non-alcoholic steatohepatitis: substudy of a phase 2, randomised placebo-controlled trial. | Wrong patient population |
| ClinicalTrials.gov. (2018). Search for Highly Specific Predictors of Response to Different Hypoglycemic Therapy for Cardiovascular Prognosis. ClinicalTrials.gov. | Wrong patient population |
| ClinicalTrials.gov. (2018). The Effects of Exenatide, a GLP-1 Agonist, on Alcohol Self-Administration in Heavy Drinkers. | Wrong follow-up duration |

| | |
|---|---|
| Ling Y.Y., Nestor L.J., Pannekoek J.N., Vanelli F., Akavarapu S., Aiano F., Kobson B., Manoharan E., Munafo M.R., Lingford-Hughes A.R., Nutt D.J., & Goldstone A.P. (2018). Do Gut Hormones attenuate eating and addictive behaviours in obesity, alcohol and nicotine dependence?. Appetite, 130, 309. https://doi.org/10.1016/j.appet.2018.05.208 | Wrong follow-up duration |
| Alkhouri, N., Herring, R., Kabler, H., Kayali, Z., Hassanein, T., Kohli, A., Huss, R., Zhu, Y., Billin, A., Damgaard, L., & et al. (2022). Safety and efficacy of combination therapy with semaglutide, cilofexor and firsocostat in patients with non-alcoholic steatohepatitis: a randomised, open-label phase II trial. https://doi.org/10.1016/j.jhep.2022.04.003 | Wrong comparator |

#### 2.11 Requests for Unpublished Data

**eTable 6 Requests for unpublished data**

| Study | Data requested | Reason for request | Outcome of request |
|---|---|---|---|
| Hendershot et al. (2025) | Unpublished smoking and alcohol outcome data | Published paper did not report all outcomes required for the meta-analysis | Authors were contacted by email. Unpublished data on alcohol outcomes were provided and included in the meta-analysis. |
| Lüthi et al. (2024) | Unpublished outcome data | Clarification and additional outcome data required for meta-analysis | Corresponding author contacted by email. Requested data were received and included in the meta-analysis. |
| Schacht et al. (2026); NCT05892432 | Unpublished alcohol outcome data | At the time of the request, trial results required for quantitative synthesis had not yet been published | Investigators were contacted on 12 February 2026. They declined to share the data while the manuscript was undergoing peer review and indicated that the data would subsequently be deposited in the NIAAA Data Archive. |
| Fogelman et al. (2025); NCT04779697 | Full numerical alcohol outcome data | Only a conference abstract was available, with insufficient numerical information for meta-analysis | Authors were contacted by email. No response or additional data is recorded in the extraction sheet; the study was therefore retained for narrative synthesis only. |
| Sarma, 2026 (Systematic review)[22] | Outcome data used in the systematic review | The values reported in the review appeared to represent baseline measurements rather than post-intervention outcome data. The review team was contacted to clarify the time point and provide the correct outcome data. | No response was received, and the review’s reported values were not used in the meta-analysis. |

#### 2.12 Risk of Bias in Included Studies

We used RoB2 to assess 41 results from 13 trials included in the meta-analyses. Overall, 2 results (4.9%) were at low risk of bias, 31 (75.6%) raised some concerns, and 8 (19.5%) were at high risk of bias (**eFigure 1**,

**eFigure 2**, **eFigure 3**). Missing outcome data were the principal concern for smoking outcomes, with 7 of 12 results judged at high risk in this domain. For alcohol outcomes, concerns commonly related to missing outcome data and outcome measurement.

**eFigure 1 Domain-level and overall risk of bias assessments for included randomised controlled trials evaluating GLP-1 receptor agonists for smoking-related outcomes**

| Study ID | Experimental | Comparator | Outcome | D1 | D2 | D3 | D4 | D5 | Overall |
|---|---|---|---|---|---|---|---|---|---|
| Ashare 2018 | Liraglutide | Placebo | Smoking cessation | ! | ! | - | + | + | - |
| Hendershot 2026 | Semaglutide | Placebo | CPD | + | ! | - | ! | + | - |
| Hendershot 2025 | Semaglutide | Placebo | CPD | + | + | - | ! | + | - |
| Klausen 2022 | Exenatide | Placebo | FTND | + | + | - | + | + | - |
| Luthi 2024 | Dulaglutide | Placebo | Smoking cessation | + | + | + | + | + | + |
| Luthi 2024 | Dulaglutide | Placebo | CPD | + | + | ! | + | + | ! |
| Luthi 2024 | Dulaglutide | Placebo | Post-cessation weight gain | + | + | + | + | + | + |
| Sass 2026 | Semaglutide | Placebo | FTND | + | + | - | + | + | - |
| Yammine 2021 | Exenatide | Placebo | Smoking cessation | + | + | ! | + | + | ! |
| Yammine 2021 | Exenatide | Placebo | Post-cessation weight gain | + | + | ! | + | + | ! |
| Klausen 2026 | Semaglutide | Placebo | FTND | + | ! | - | ! | + | - |
| Klausen 2026 | Semaglutide | Placebo | CPD | + | ! | - | ! | + | - |

**eFigure 2 Domain-level and overall risk of bias assessments for included randomised controlled trials evaluating GLP-1 receptor agonists for alcohol-related outcomes**

| Study ID | Experimental | Comparator | Outcome | D1 | D2 | D3 | D4 | D5 | Overall |
|---|---|---|---|---|---|---|---|---|---|
| Armstrong 2016 | Liraglutide | Placebo | Alcohol (g/d) | + | + | - | + | + | - |
| Hendershot 2025 | Semaglutide | Placebo | Alcohol (g/d) | + | + | + | ! | + | ! |
| Hendershot 2025 | Semaglutide | Placebo | Heavy drinking days | + | + | + | ! | + | ! |
| Hendershot 2025 | Semaglutide | Placebo | Drinks per drinking day | + | + | + | ! | + | ! |
| Hendershot 2025 | Semaglutide | Placebo | Drinks per calendar day | + | + | + | ! | + | ! |
| Hendershot 2025 | Semaglutide | Placebo | Drinking days | + | + | + | ! | + | ! |
| Hendershot 2025 | Semaglutide | Placebo | Days w/o alcohol | + | + | + | ! | + | ! |
| Klausen 2022 | Exenatide | Placebo | Heavy drinking days | + | + | ! | + | + | ! |
| Klausen 2022 | Exenatide | Placebo | AUDIT | + | + | ! | + | + | ! |
| Klausen 2022 | Exenatide | Placebo | PACS | + | + | ! | + | + | ! |
| Klausen 2022 | Exenatide | Placebo | Days w/o alcohol | + | + | ! | + | + | ! |
| Klausen 2022 | Exenatide | Placebo | Alcohol (g/d) | + | + | ! | + | + | ! |
| Larsen 2017 | Liraglutide | Placebo | AUDIT | + | + | ! | + | + | ! |
| Larsen 2017 | Liraglutide | Placebo | Alcohol (d/w) | + | + | ! | + | + | ! |
| Luthi 2024 | Dulaglutide | Placebo | Alcohol (d/w) | + | + | ! | + | + | ! |
| Miras 2019 | Liraglutide | Placebo | AUDIT | + | + | ! | + | + | ! |
| Sass 2026 | Semaglutide | Placebo | AUDIT | + | + | ! | + | + | ! |
| Vedtofte 2020 | Liraglutide | Placebo | Alcohol (d/w) | + | + | ! | + | + | ! |
| Schacht 2026 | Semaglutide | Placebo | PACS | + | + | ! | ! | ! | ! |
| Schacht 2026 | Semaglutide | Placebo | Drinks per drinking day | + | + | ! | ! | ! | ! |
| Schacht 2026 | Semaglutide | Placebo | Drinks per calendar day | + | + | ! | ! | ! | ! |
| Schacht 2026 | Semaglutide | Placebo | Drinking days | + | + | ! | ! | ! | ! |
| Schacht 2026 | Semaglutide | Placebo | Heavy drinking days | + | + | ! | ! | ! | ! |
| Klausen 2026 | Semaglutide | Placebo | PACS | + | ! | ! | ! | + | ! |
| Klausen 2026 | Semaglutide | Placebo | AUDIT | + | ! | ! | ! | + | ! |
| Klausen 2026 | Semaglutide | Placebo | Drinks per drinking day | + | ! | ! | ! | + | ! |
| Klausen 2026 | Semaglutide | Placebo | Heavy drinking days | + | ! | + | ! | + | ! |
| Klausen 2026 | Semaglutide | Placebo | Alcohol (g/d) | + | ! | + | ! | + | ! |
| Klausen 2026 | Semaglutide | Placebo | Days w/o alcohol | + | ! | + | ! | + | ! |

**eFigure 3 Distribution of RoB 2 judgements across bias domains and overall risk of bias for smoking- and alcohol-related outcomes. Bars show unweighted percentages of assessed results.**

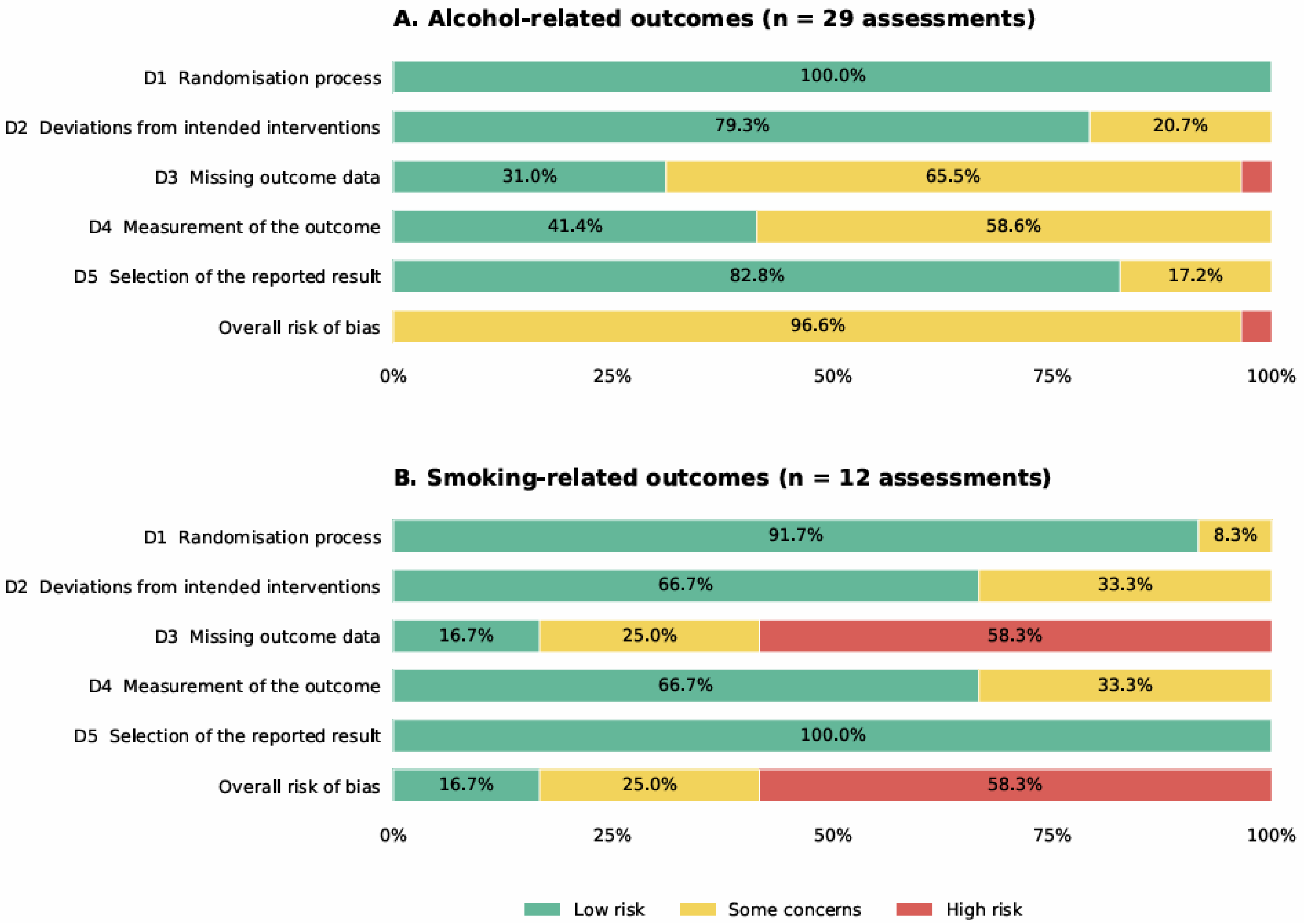

Unweighted proportions: each study-outcome assessment contributes equally. Studies with multiple outcomes appear more than once. Bars do not represent meta-analysis weights or proportions of unique trials.

##### 2.13 Certainty of Evidence (GRADE)

Certainty of evidence ranged from very low to moderate (**eTable 7** and **eTable 8**). For smoking outcomes, certainty was low for cessation, very low for cigarettes smoked per day and nicotine dependence, and moderate for post-cessation weight. For alcohol outcomes, certainty was low for drinks per drinking day, drinks per week and days without alcohol consumption, and very low for all other assessed outcomes.

#### eTable 7 Summary of findings and GRADE certainty of evidence for GLP-1 receptor agonists versus placebo on smoking-related outcomes

| **Certainty assessment** | | | | | | | **№ of patients** | | **Effect** | | **Certainty** |
|---|---|---|---|---|---|---|---|---|---|---|---|
| **№ of studies** | **Study design** | **Risk of bias** | **Inconsistency** | **Indirectness** | **Imprecision** | **Other considerations** | **GLP-1s** | **Placebo** | **Relative (95%CI)** | **Absolute (95%CI)** | |
| **Smoking cessation** | | | | | | | | | | | |
| 3 | randomised trials | serious[a] | not serious | not serious | serious[b] | none | 58/190 (30.5%) | 48/187 (25.7%) | **OR 1.39** (0.77 to 2.53) | 68 more per 1,000 (from 49 fewer to 210 more) | ⨁⨁◯◯ Low[a,b] |
| **Cigarettes per day** | | | | | | | | | | | |
| 4 | randomised trials | very serious[c] | serious[d] | not serious | serious[e] | none | 200 | 200 | - | SMD **0.21 SD lower** (0.44 lower to 0.02 higher) | ⨁◯◯◯ Very low[c,d,e] |
| **Nicotine dependence** | | | | | | | | | | | |
| 3 | randomised trials | very serious[f] | serious[g] | not serious | very serious[h] | none | 105 | 103 | - | SMD **0.08 SD lower** (0.6 lower to 0.44 higher) | ⨁◯◯◯ Very low[f,g,h] |
| **Post-cessation weight** | | | | | | | | | | | |
| 2 | randomised trials | not serious[i] | not serious | not serious | serious[j] | none | 52 | 52 | - | SMD **0.27 SD lower** (0.58 lower to 0.05 higher) | ⨁⨁⨁◯ Moderate[i,j] |

Explanations: a. Downgraded by one level for risk of bias. One of three studies was overall high risk of bias, one had some concerns and one was low risk across all domains. b. Downgraded by one level for imprecision. OIS for a dichotomous outcome (n=586) not met. The 95%CI (0.77 to 2.53) crosses the null and the threshold for clinically important benefit, with the lower bound just inside the threshold for harm. c. Downgraded by two levels for risk of bias. Three of four studies were overall high risk of bias, driven by missing outcome data. d. Downgraded by one level for inconsistency. I2 (61%) substantial, point estimates range from near null to moderate effect; direction of effect broadly consistent but magnitude varies. e. Downgraded by one level for imprecision. OIS (n = 786) not met, and CI crosses null. f. Downgraded by two levels for risk of bias. All three studies were overall high risk of bias, driven by missing outcome data. g. Downgraded by one level for inconsistency. I2 (63%) substantial; point estimates diverge in direction and some CIs overlap. h. Downgraded by two levels for imprecision. OIS (n = 786) not met, CI crosses thresholds for clinically important benefit and clinically important harm. i. Not downgraded. One of two studies was low risk of bias across all domains - other had some concerns in a single domain. Neither was rated as high risk overall. j. Downgraded by one level for imprecision. OIS (n=786) not met (only 2 studies). CI (-0.58, 0.05) crosses the null, ranging from a clinically important benefit to no effect.

#### eTable 8 Summary of findings and GRADE certainty of evidence for GLP-1 receptor agonists versus placebo on alcohol-related outcomes

| Certainty assessment | | | | | | | № of patients | | Effect | | Certainty |
|---|---|---|---|---|---|---|---|---|---|---|---|
| № of studies | Study design | Risk of bias | Inconsistency | Indirectness | Imprecision | Other considerations | GLP-1s | Placebo | Relative (95%CI) | Absolute (95%CI) | |
| **Alcohol grams daily** | | | | | | | | | | | |
| 4 | randomised trials | serious[a] | serious[b] | not serious | serious[c] | none | 163 | 165 | - | SMD 0.3 SD lower (0.63 lower to 0.04 higher) | ⨁◯◯◯ Very low[a,b,c] |
| **Alcohol dependence** | | | | | | | | | | | |
| 5 | randomised trials | serious[d] | serious[e] | not serious | very serious[f] | none | 244 | 227 | - | SMD 0.03 SD lower (0.29 lower to 0.23 higher) | ⨁◯◯◯ Very low[d,e,f] |
| **Alcohol craving** | | | | | | | | | | | |
| 3 | randomised trials | serious[g] | serious[h] | not serious | very serious[i] | none | 140 | 145 | - | SMD 0.33 SD lower (0.85 lower to 0.18 higher) | ⨁◯◯◯ Very low[g,h,i] |
| **Drinks per drinking day** | | | | | | | | | | | |
| 3 | randomised trials | serious[g] | not serious | not serious | serious[j] | none | 102 | 104 | - | SMD 0.59 SD lower (0.86 lower to 0.32 lower) | ⨁⨁◯◯ Low[g,j] |
| **Drinks per calendar day** | | | | | | | | | | | |
| 2 | randomised trials | serious[g] | not serious | not serious | very serious[k] | none | 48 | 50 | - | SMD 0.48 SD lower (0.87 lower to 0.09 lower) | ⨁◯◯◯ Very low[g,k] |
| **Drinks per week** | | | | | | | | | | | |
| 3 | randomised trials | serious[l] | not serious | not serious | serious[m] | none | 160 | 170 | - | SMD 0.12 SD lower (0.29 lower to 0.05 higher) | ⨁⨁◯◯ Low[l,m] |
| **Heavy drinking days** | | | | | | | | | | | |
| 4 | randomised trials | serious[n] | serious[o] | not serious | very serious[p] | none | 124 | 125 | - | SMD 0.28 SD lower (0.71 lower to 0.15 higher) | ⨁◯◯◯ Very low[n,o,p] |
| **Days without alcohol consumption** | | | | | | | | | | | |
| 3 | randomised trials | serious[g] | not serious | not serious | serious[q] | none | 142 | 135 | - | SMD 0.35 SD higher (0.12 higher to 0.59 higher) | ⨁⨁◯◯ Low[g,q] |

| | | | | | | | | | | | |
|---|---|---|---|---|---|---|---|---|---|---|---|
| **Drinking days** | | | | | | | | | | | |
| 2 | randomised trials | serious[g] | not serious | not serious | very serious[k] | none | 48 | 50 | - | SMD 0.06 SD lower (0.18 lower to 0.05 higher) | ⊕◯◯◯ Very low[g,k] |

Explanations: a. Downgraded by one level for risk of bias. One of four studies was overall high risk of bias due to missing outcome data; the remaining three studies had some concerns. No study was low risk across all domains.b. Downgraded by one level for inconsistency. Point estimates vary in direction of effect, moderate statistical heterogeneity (I2 = 62%). c. Downgraded by one level for imprecision. Meta-analysis did not meet OIS (n = 786) for a threshold SMD of 0.2 and 95%CI (-0.63, 0.04) crosses null ranging from clinically important benefit to no effect. d. Downgraded by one level for risk of bias. All five studies were judged overall as some concerns, primarily due to missing outcome data; one had additional concerns in deviations from intended interventions and outcome measurement. No study was rated low or high risk of bias overall. e. Downgraded by one level for inconsistency. Heterogeneity was substantial ($I^2$=65%) with point estimates varying in direction across studies. f. Downgraded by two levels for imprecision. Meta-analysis did not meet OIS (n = 786), and the 95%CI (-0.29, 0.23) crossed thresholds for both clinically important benefit and harm. g. Downgraded by one level for risk of bias. All studies were judged overall as some concerns. No study was rated low or high risk of bias overall. h. Downgraded by one level for inconsistency. Heterogeneity was substantial (I2 = 78%), with point estimates varying in direction across studies; some CIs overlap. i. Downgraded by two levels for imprecision. Meta-analysis did not meet OIS (n = 786) and the 95%CI (-0.85 to 0.18) crossed thresholds for clinically important benefit and harm. j. Sample size (n=206) did not meet OIS (n=786), the 95%CI (-0.86, -0.32) excludes the null and the threshold for harm, and lay beyond the 0.2 threshold for a clinically important benefit. k. Sample size (<100 participants) extremely small - so reduced by two levels. l. Downgraded by one level for risk of bias. All studies were judged overall as some concerns. No study was rated as low or high risk of bias overall, m. Downgraded by one level for imprecision. Meta-analysis (n=330) did not meet OIS (n=786), and the 95%CI (-0.29, 0.05) crossed the null, ranging from a clinically important benefit to no effect. n. Downgraded by one level for risk of bias. All studies were judged overall as some concerns. No study was rated low/high risk of bias overall. o. Downgraded by one level for inconsistency. Heterogeneity was substantial (I2 = 80%), with point estimates varying in direction across studies; some CIs overlap. p. Downgraded by two levels for imprecision. Meta-analysis did not meet OIS (n = 786) and the 95%CI (-0.71, 0.15) crossed thresholds for clinically important benefit/null. q. Downgraded by one level. Meta-analysis did not meet OIS (n = 786), the 95%CI (0.12, 0.59) ranged minimal to a large effect.

##### 2.14 Funnel Plots and Small-Study Effects

The funnel plots contained only 2 to 5 studies per outcome, making visual assessment of asymmetry unreliable (**eFigure 4** and **eFigure 5**). Exploratory Egger's tests showed little evidence of funnel-plot asymmetry for most outcomes (**eTable 9**). The exception was nicotine dependence measured by the FTND (p=0.04), but this test was based on only 3 studies and should not be interpreted as evidence of publication bias. Every analysis included fewer than the 10 studies recommended for reliable testing, so the test results are presented for illustration only[2]. We did not run Egger's tests for outcomes with only 2 studies.

**eFigure 4 Funnel plots for smoking outcomes in randomised trials of GLP-1 receptor agonists***

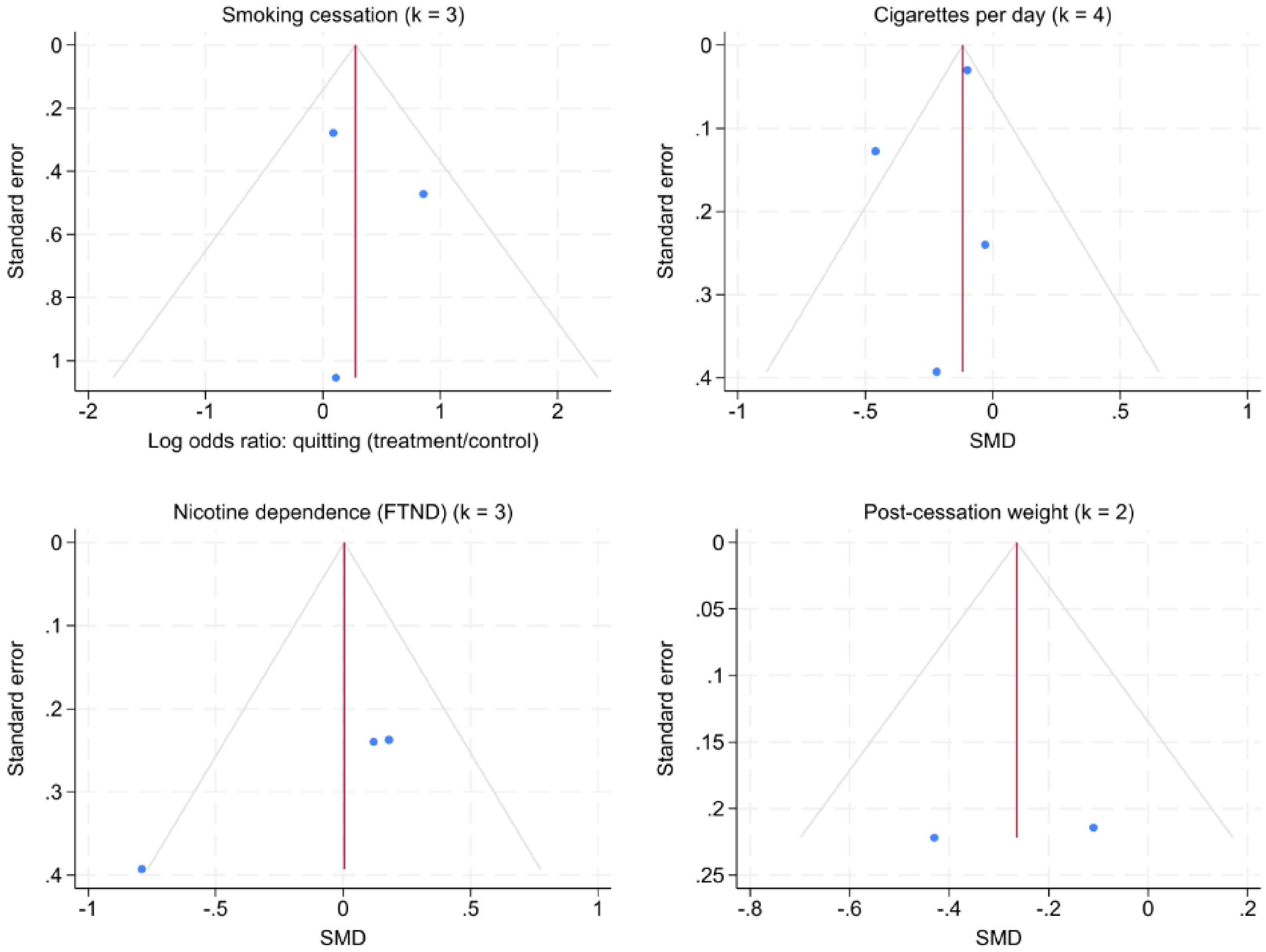

*Smoking cessation: log odds ratio; other outcomes: SMD. Figure note: Few studies contributed to each outcome. The plots are descriptive; apparent asymmetry does not establish publication bias.

#### eFigure 5 Funnel plots for alcohol outcomes in randomised trials of GLP-1 receptor agonists*

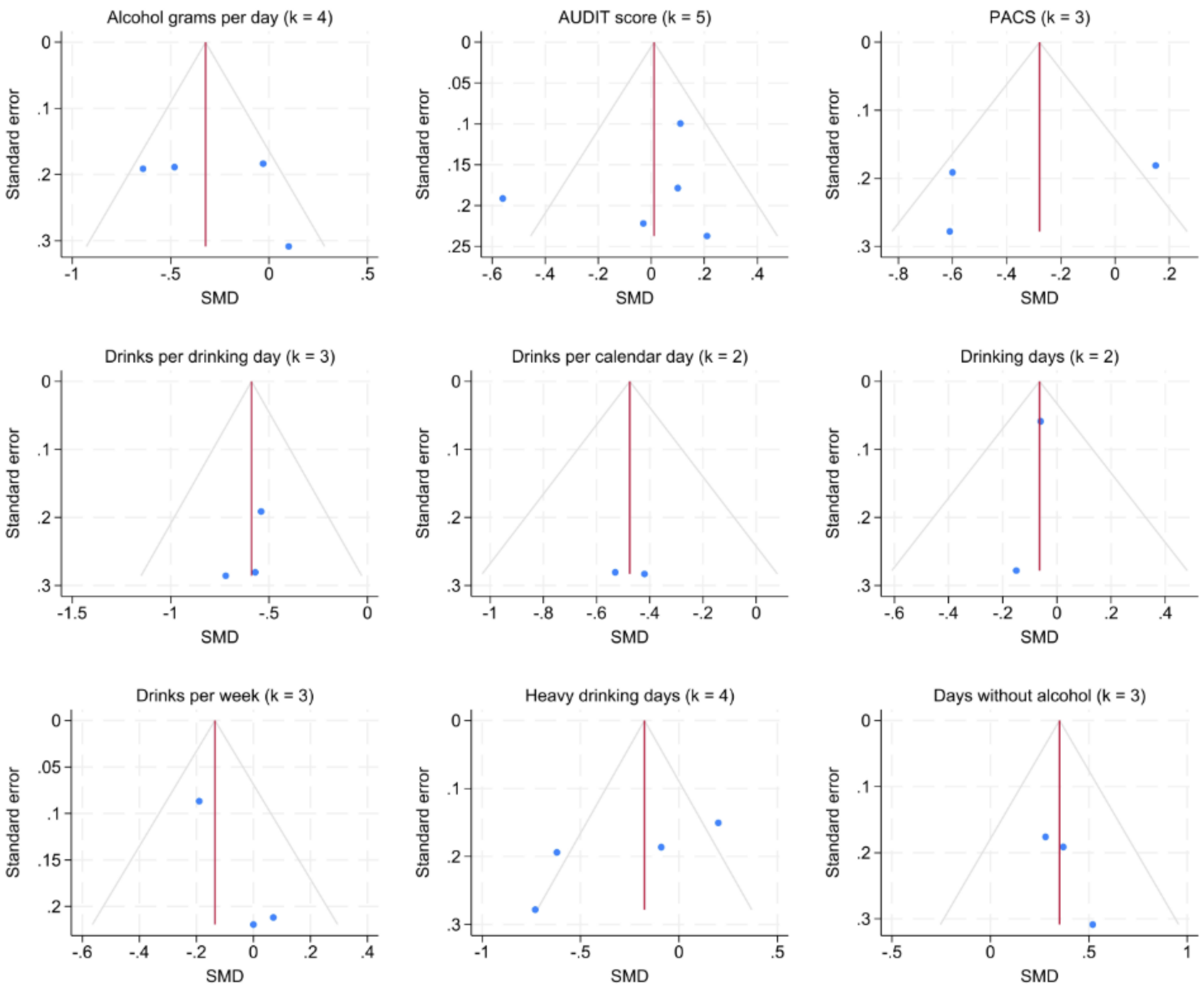

*Figure note: Few studies contributed to each outcome. The plots are descriptive; apparent asymmetry does not establish publication bias.

#### eTable 9 Exploratory tests for small-study effects across smoking and alcohol outcomes

| Outcome | k | Egger's test | P value |
|---|---|---|---|
| Smoking cessation | 3 | 0.39 | 0.77 |
| Cigarettes per day (CPD) | 4 | -0.79 | 0.51 |
| Fagerström Test for Nicotine Dependence (FTND) | 3 | -16.72 | 0.04 |
| Post-cessation weight | 2 | - | - |
| Daily alcohol consumption (grams) | 4 | 0.71 | 0.55 |
| Alcohol Use Disorders Identification Test (AUDIT) | 5 | -0.61 | 0.59 |
| Penn Alcohol Craving Scale (PACS) | 3 | -0.63 | 0.64 |
| Drinks per drinking day | 3 | -1.08 | 0.48 |
| Drinks per calendar day | 2 | - | - |
| Drinking days | 2 | - | - |
| Drinks per week | 3 | 4.69 | 0.13 |
| Heavy drinking days | 4 | -2.10 | 0.17 |
| Days without alcohol consumption | 3 | 2.56 | 0.24 |
| *Note:* Egger's test was not run where (k=2). All tests were exploratory because each outcome included fewer than 10 studies; the FTND *p* value should not be interpreted as evidence of publication bias. | | | |

#### *3. REFERENCES*

1 Follmann, D., Elliott, P., Suh, I. & Cutler, J. Variance imputation for overviews of clinical trials with continuous response. *Journal of Clinical Epidemiology* **45**, 769-773 (1992). https://doi.org/10.1016/0895-4356(92)90054-Q

2 Higgins, J. *et al. Cochrane Handbook for Systematic Reviews of Interventions*. Vol. 6.5 (Cochrane, 2024).

3 Daly, C., Welton, N. J., Dias, S., Anwer, S. & Ades, A. Meta-Analysis of Continuous Outcomes. Guideline Methodology Document 2 (Version 1). (University of Bristol, 2021).

4 Paterson, C. *et al.* Efficacy and safety of pharmacological interventions for the treatment of cocaine use disorder: a systematic review and network meta-analysis. *medRxiv*, 2025.2012.2010.25341965 (2025). https://doi.org/10.64898/2025.12.10.25341965

5 Taylor, G. *et al.* Smoking cessation for improving mental health. *Cochrane Database*, CD013522 (2021). https://doi.org/10.1002/14651858.CD013522.pub2

6 StataCorp LLC. *Stata 19 Meta-Analysis Reference Manual*. Vol. Release 19 (Stata Press, 2025).

7 Taylor, G. *et al.* Incretin mimetics for treatment of tobacco and alcohol dependence: a protocol for a systematic review and meta-analysis of RCTs and experiments (ID: CRD420251155420). *PROSPERO* (2025).

8 Klausen, M. K. *et al.* Exenatide once weekly for alcohol use disorder investigated in a randomised, placebo-controlled clinical trial. *JCI Insight* **7** (2022). https://doi.org/10.1172/jci.insight.159863

9 Larsen, J. R. *et al.* Effect of Liraglutide Treatment on Prediabetes and Overweight or Obesity in Clozapine- or Olanzapine-Treated Patients With Schizophrenia Spectrum Disorder: A Randomised Clinical Trial. *JAMA Psychiatry* **74**, 719-728 (2017). https://doi.org/10.1001/jamapsychiatry.2017.1220

10 Sass, M. R. *et al.* Semaglutide and Early-Stage Metabolic Abnormalities in Individuals With Schizophrenia Spectrum Disorders: A Randomised Clinical Trial. *JAMA Psychiatry* **83**, 128-138 (2026). https://doi.org/10.1001/jamapsychiatry.2025.3639

11 Vedtofte, L. *et al.* One Year's Treatment with the Glucagon-Like Peptide 1 Receptor Agonist Liraglutide Decreases Hepatic Fat Content in Women with Nonalcoholic Fatty Liver Disease and Prior Gestational Diabetes Mellitus in a Randomised, Placebo-Controlled Trial. *J Clin Med* **9** (2020). https://doi.org/10.3390/jcm9103213

12 Klausen, M. K. *et al.* Once-weekly semaglutide versus placebo in patients with alcohol use disorder and comorbid obesity: a randomised, double-blind, placebo-controlled trial. *The Lancet* **407**, 1687-1698 (2026). https://doi.org/10.1016/S0140-6736(26)00305-3

13 Ashare, R. Daily Liraglutide for Nicotine Dependence (NCT03712098). (ClinicalTrials.gov, 2018).

14 Fogelman, N. *et al.* Semaglutide compared to placebo treatment decreases alcohol consumption and food craving over 12-weeks and a 1-month follow up. *Alcohol-Clinical and Experimental Research* **49** (2025).

15 Hendershot, C. S. *et al.* Once-Weekly Semaglutide in Adults With Alcohol Use Disorder: A Randomised Clinical Trial. *JAMA Psychiatry* **82**, 396–404 (2025). https://doi.org/10.1001/jamapsychiatry.2024.4789
16 Hendershot, C. S. *et al.* Once-Weekly Semaglutide in Adults With Daily Cigarette Use: A Randomised Clinical Trial. *JAMA Network Open* **9**, e2614898 (2026). https://doi.org/10.1001/jamanetworkopen.2026.14898
17 Yammine, L. *et al.* Exenatide Adjunct to Nicotine Patch Facilitates Smoking Cessation and May Reduce Post-Cessation Weight Gain: A Pilot Randomised Controlled Trial. *Nicotine and Tobacco Research* **23**, 1682EP-1690 (2021). https://doi.org/10.1093/ntr/ntab066
18 Schacht, J. P. S., Joseph T; Raymond, Kristen; Shelton, Robert. Oral Semaglutide for Alcohol Use Disorder: A Randomised Clinical Trial. *The American Journal of Psychiatry* **1**, 636-645 (2026). https://doi.org/10.1176/appi.ajp.20260003
19 Armstrong, M. J. *et al.* Liraglutide safety and efficacy in patients with non-alcoholic steatohepatitis (LEAN): a multicentre, double-blind, randomised, placebo-controlled phase 2 study. *Lancet* **387**, 679-690 (2016). https://doi.org/10.1016/s0140-6736(15)00803-x
20 Miras, A. D. *et al.* Adjunctive liraglutide treatment in patients with persistent or recurrent type 2 diabetes after metabolic surgery (GRAVITAS): a randomised, double-blind, placebo-controlled trial. *Lancet Diabetes Endocrinol* **7**, 549-559 (2019). https://doi.org/10.1016/s2213-8587(19)30157-3
21 Luthi, H. *et al.* Effect of dulaglutide in promoting abstinence during smoking cessation: 12-month follow-up of a single-centre, randomised, double-blind, placebo-controlled, parallel group trial. *eClinicalMedicine* **68**, 102429 (2024). https://doi.org/10.1016/j.eclinm.2024.102429
22 Sarma, I. *et al.* A Systematic Review and Meta-Analysis Evaluating the Role of GLP-1 Receptor Agonists in Substance Use Disorders. *Cureus* **18** (2026/06/28). https://doi.org/10.7759/cureus.111641
23 Hendershot, C. S. Effects of Semaglutide on Nicotine Intake and Smoking LapseA1 (NCT05530577). *clinicaltrials.gov* (2022).
